# *OCT1* Variants Are Associated with Metformin Clearance and Gluconeogenesis: Mechanistic Insights for Youth-Onset Type 2 Diabetes in the MIGHTY Study

**DOI:** 10.64898/2026.05.27.26354152

**Authors:** Samson L. Cantor, Yi Zeng, Faith S. Davis, Sophia B. Glaros, Natalie A. Macheret, Ila N. Kacker, Noemi Malandrino, Lilian Mabundo, Oluwatobi T. Arisa, Adebowale A. Adeyemo, Hongyi Cai, Amber B. Courville, Eileen L. Shouppe, Mary F. Walter, Peter J. Walter, Charles N. Rotimi, William D. Figg, Amy R. Bentley, Stephanie T. Chung

**Affiliations:** National Institutes of Health, National Institute of Diabetes and Digestive Disorders and Kidney Disease, Section on Pediatric Diabetes, Obesity, and Metabolism, Bethesda, MD, US; National Institutes of Health, Clinical Center, Clinical Pharmacology Laboratory Bethesda, MD, US; National Institutes of Health, National Human Genome Research Institute, Center for Research on Genomics and Global Health, Bethesda, MD, US

**Keywords:** glycemia, pediatric diabetes, pharmacokinetics, pharmacogenetics, polymorphism, *SLC22A1*, *TCF7L2*

## Abstract

**Aims/Hypothesis:** Behavioral and phenotypic characteristics do not fully explain variability in African Americans with youth-onset type 2 diabetes (Y-T2D) treated with metformin with or without liraglutide. We hypothesized that biological heterogeneity, including genetic variation in the metformin transporter *OCT1,* influences metformin pharmacokinetics and hepatic glucose flux. Therefore, we sought to characterize metformin pharmacokinetics in Y-T2D and evaluate genetic variants known to modulate metformin efficacy in adults to determine the mechanisms underlying variation in treatment response.

**Methods:** We evaluated genetic variants related to metformin transport and mechanisms of action in 30 Y-T2D using a candidate-gene approach to evaluate the association of pharmacogenetic variants with fasting glucose and gluconeogenesis. In a subset of Y-T2D randomized to 3 months of metformin (n=11) or metformin and liraglutide (n=8), we constructed a metformin population pharmacokinetic model and evaluated gene variant associations.

**Results:** A one-compartment first-order absorption and elimination pharmacokinetic model provided the optimal fit. Metformin pharmacokinetic parameters were similar by group and not related to glycemia. The rs628031_*OCT1* A allele was associated with greater metformin clearance. The rs622342_*OCT1* C allele was associated with lower post-treatment fractional gluconeogenesis (β [95% CI] = -8.8 [-14.13, -3.47] %, Adjusted R^2^ = 0.56, *P* = 0.003). The rs7903146_*TCF7L2* T allele was associated with greater reductions in fasting glucose among those treated with metformin + liraglutide (β = -1.32 [-2.42, -0.22] mmol/L, Adjusted R^2^ = 0.8, *P*<0.002), but baseline glucose and gluconeogenesis (*P*<0.0001) were the strongest predictors of post-treatment glycemia.

**Conclusion/interpretation:** In Y-T2D, *OCT1* gene variants rs628031 and rs622342 were associated with metformin clearance and gluconeogenesis, respectively. *TCF7L2* variant rs7903146 may contribute to differences in glycemic response in youth treated with metformin and liraglutide. These findings suggest genetic variants may be important for understanding variable metformin response in Y-T2D.

**RESEARCH IN CONTEXT:** *What is already known about this subject?:*

- Metformin is the first-line therapeutic approach for management of youth-onset type-2 diabetes (Y-T2D); however, 65% of African American (AA) youth require additional medication within 2 years, and the biological drivers of variability remain unclear
- Y-T2D has a distinct pathophysiology from adult-onset T2D, characterized by rapid beta-cell failure, high rates of gluconeogenesis, and glomerular hyperfiltration that may influence metformin action and clearance, especially among AA Y-T2D.
- Genetic variants in the OCT1 (*SLC22A1*) metformin transporter have been associated with metformin clearance and glycemic response in adults, but there are no pharmacokinetic-pharmacogenetic models of metformin in Y-T2D

*What is the key question?:*

- Among AA Y-T2D, how do genetic variants associated with adult-onset T2D affect metformin pharmacokinetic profiles and glycemic response?

*What are the new findings?:*

- Metformin pharmacokinetics in AA Y-T2D were best described by a one-compartment first-order absorption and elimination model
- The *OCT1* variant rs628031 A allele was associated with ∼40% greater metformin clearance
- The *OCT1* variant rs622342 C allele was associated with ∼8% lower rates of fractional gluconeogenesis after treatment

*How might this impact on clinical practice in the foreseeable future?:*

- Identifying *OCT1* variants that modulate gluconeogenic flux, together with a novel pharmacokinetic profile, could help reshape evaluation of metformin response in Y-T2D and facilitate mechanistically informed, genotype-guided strategies to distinguish patients who can be safely managed on metformin from those who need early combination therapy

## 1. Introduction

Metformin is the recommended first-line therapy and the most widely prescribed diabetes medication in youth-onset type 2 diabetes (Y-T2D) [1]. However, metformin efficacy—absolute reductions in Hemoglobin A1c (HbA1c) or fasting glucose—is highly variable, especially in African American Y-T2D, 65% of whom require additional medications within 2 years of starting treatment [2]. The biological, genetic, and contextual factors contributing to reduced metformin responsiveness in Y-T2D are an ongoing field of study and were the focus of the Metformin Influences Gut Hormone in Youth (MIGHTY) group of studies. The MIGHTY studies evaluated metformin mechanisms of action and disease pathogenesis in African American Y-T2D at increased risk for treatment failure. Unique characteristics of Y-T2D, including high rates of gluconeogenesis and rapid beta-cell failure, were early and prominent pathogenic features that exceeded the capacity of metformin’s glucose lowering effect [3, 4]. Short-term treatment with metformin monotherapy or in combination with liraglutide, a glucagon-like peptide-1 receptor agonist (GLP-1RA), was insufficient for mitigating high rates of gluconeogenesis and whole-body lipolysis in African American Y-T2D [5]. Furthermore, heterogeneity in reducing rates of gluconeogenesis and glycemia was not readily explained by common demographic or behavioral characteristics, such as duration of diabetes or medication adherence. Combining metformin with liraglutide reduced HbA1c but did not lower rates of gluconeogenesis, a primary contributor to fasting glucose concentrations.

Heritability of metformin’s glycemic effect has been estimated at 12-34% [6, 7], suggesting a genetic component may underlie treatment variability. Therefore, we hypothesized that variants in genes encoding metformin transport enzymes may contribute to the observed phenomenon; however, no studies have evaluated pharmacokinetic-pharmacodynamic-pharmacogenomic mechanisms in Y-T2D. To date, only one genome-wide association study of metformin response has been conducted in Y-T2D [8]. The TODAY study of 506 Y-T2D, which included 37% African American participants, showed initial evidence of genetic variation in metformin response, although no associations achieved genome-wide significance. Exploring genetic variation as a key predictor of metformin failure in Y-T2D is also supported because contextual factors such as gastrointestinal adverse events [5, 9], phenotypic (age, sex, body mass index [10]), or behavioral characteristics (medication nonadherence [2, 5]) have not explained variability in metformin’s glucose lowering effect in Y-T2D.

Metformin’s uptake and excretion are entirely dependent on solute carriers and shuttle proteins, Figure 1 [11, 12]. Gene variants encoding solute carriers, such as the organic cation transporters (OCT) and multidrug and toxin extrusion proteins (MATE), are linked to metformin clearance, reduction in HbA1c, and intolerance in adults in some [13–17], but not all studies [18, 19]. The strongest evidence for variants affecting metformin transport and efficacy exists for *OCT1* (also known as *SLC22A1*), though genetic polymorphisms in *OCT2* (*SLC22A2*)*, GLUT2* (*SLC2A2*)*, MATE1* (*SLC47A1*), *MATE2* (*SLC47A2*), and *ATM* (ataxia telangiectasia mutated) may also be important mechanistically [20–22]. Importantly, these studies have all been conducted in adults with T2D, and there are unique characteristics of Y-T2D compared to adult-onset T2D that warrant separate study of Y-T2D. For example, recent data indicate glomerular hyperfiltration is prevalent in Y-T2D [23] and could modulate drug clearance, motivating the need for a pharmacokinetic-pharmacogenetic (PK-PG) model of metformin response in youth.

**Figure 1:**
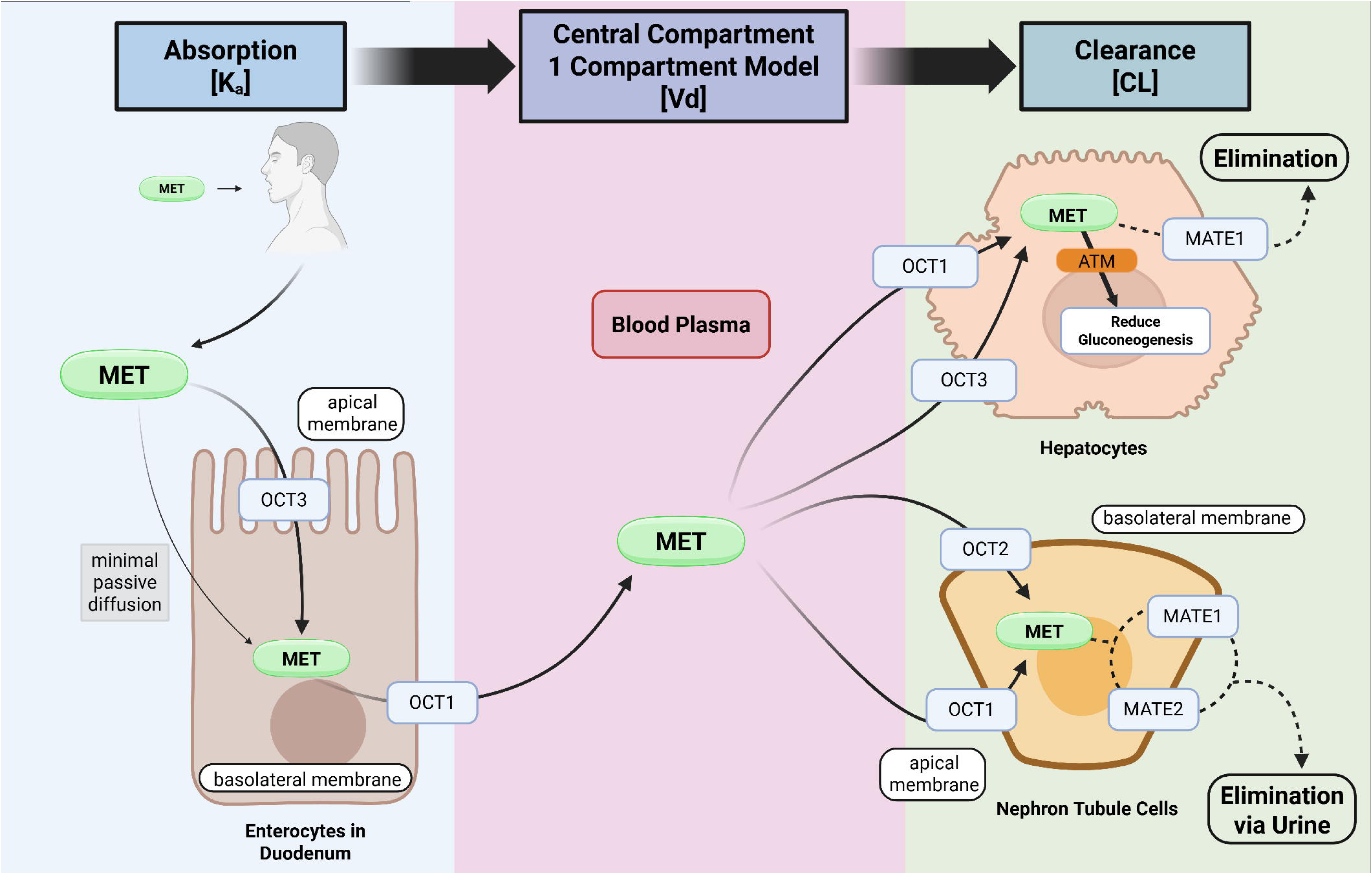
Mechanistic model of metformin across pharmacokinetic stages. This figure is a schematic representation of metformin (MET) absorption, distribution, and clearance within a one-compartment pharmacokinetic framework. The organic cation transporter proteins 1-3 (OCT1, OCT2, OCT3), are highly implicated in this process. During absorption (Ka, absorption rate constant), metformin (MET) is taken up from the intestinal lumen into enterocytes in the duodenum, with minimal passive diffusion due to its basic, hydrophilic nature. OCT1 subsequently transports metformin across the basolateral membrane into the blood plasma, the central compartment. In the central compartment, which represents the volume of distribution (Vd), metformin uptake by hepatocytes occurs via OCT1 and OCT3, activating ATM and the downstream AMP-activated protein kinase pathway to reduce hepatic gluconeogenesis. Clearance (CL) occurs through MATE1-mediated transport in the hepatocytes alongside renal elimination. In the nephron tubule cells, uptake occurs via OCT 1 and OCT2 in the basolateral membrane and efflux via multidrug and toxin extrusion transporters 1 and 2 (MATE1 and MATE2) on the apical membrane, leading to urinary excretion. Created with BioRender.com [12]. Abbreviations: Ka, absorption rate constant; Vd, volume of distribution; CL, clearance; OCT, organic cation transporter; MATE, multidrug and toxin extrusion transporter; ATM, ataxia telangiectasia mutated; MET, metformin.

Additionally, existing PK-PG models in adult diabetes may not adequately capture the rapid disease progression observed in Y-T2D. Only one PK-PG model of metformin response is available in youth; however, this study included youth with obesity but without diabetes [19]. Drug-drug interactions may also influence treatment efficacy, and no studies have evaluated genotype interactions among Y-T2D users of both metformin and liraglutide, although combined therapy is common. Data on genetic determinants of liraglutide response are just emerging in adults [24–26]. To address these knowledge gaps, our overall goals are to help differentiate young people who may be safely and effectively managed on metformin from those who require early combination therapy.

This analysis was designed to integrate known genetic determinants of metformin transport and liraglutide response with glucose metabolism in African American youth to help understand and uncover therapeutic targets that could be optimized for high-risk youth. We explored the mechanisms influencing metformin action by employing a hypothesis-generating, candidate-gene approach that integrated population-specific pharmacokinetic modeling with deep metabolic phenotyping. We developed a multi-locus genetic risk score for metformin response, drawing from established associations in the literature and focusing on variants with a high minor allele frequency (>20%) among individuals of African American ancestry. The study objectives were to: (1) conduct a pharmacogenetic candidate gene analysis of single nucleotide polymorphisms previously associated with metformin response, (2) develop a population pharmacokinetic-pharmacodynamic model, and (3) evaluate predictors of fasting glucose and gluconeogenesis after 3 months of treatment with metformin with or without liraglutide.

## 2. Methods

### 2.1 Study design

This study was conducted at the Metabolic Clinical Research Unit at the NIH Clinical Center between 2017 and 2022 [5, 27]. The primary study objectives compared the change in rates of gluconeogenesis and beta-cell function [5]. This exploratory analysis was designed to evaluate genetic and pharmacologic factors that may influence short-term glycemic response to metformin and liraglutide in Y-T2D.

### 2.2 Participants

Participants included in this PK-PD-PG analysis had complete genetic data (n=30), of which a subset were randomized into a two-arm parallel trial of MET (n=14) and MET+LIRA (n=11), and 19 had complete data for PK-PD-PG modeling (Supplemental Figure 1). All youth were aged 12-25 years, diagnosed with Y-T2D by the American Diabetes Association criteria within ≤5 years [28], had baseline HbA1c ≤9% (≤75mmol/mol), were negative for diabetes autoantibodies (glutamic acid decarboxylase-65 and insulinoma-associated protein-2 autoantibodies), had never received GLP1-RAs, and were not on insulin treatment in the preceding 3 months.

### 2.3 Trial procedures and medication titration

Participants were evaluated during two inpatient visits—once at baseline after a 5–7-day medication washout period for those previously on metformin therapy and again after receiving 3 months of MET or MET+LIRA (Supplemental Figure 2a). Randomization to treatment group occurred at the baseline visit, and participants started oral immediate release MET (500mg tablets) with or without LIRA (0.6mg subcutaneous injection daily). A standard protocol was used for medication titration; the maximum titrated dose over 3 weeks was MET 1000mg twice daily and LIRA 1.8mg daily for 12 ± 2 weeks. Patients with gastrointestinal side effects or fasting blood glucose <75 mg/dL (<4.16 mmol/L) on 2 or more consecutive days were maintained at their maximum tolerable dose according to a standard protocol (Supplemental materials). Adherence was assessed following intervention via pill and/or pen count and serum metformin and/or liraglutide concentration measured by HPLC-MS/MS. Lean body mass was assessed with dual-energy X-ray absorptiometry (GE Healthcare, Madison, WI, USA).

The study procedures have been previously described [5]; all procedures, including energy needs, were standardized during study visits for all participants. Standard-of-care recommendations were provided for free-living diet and physical activity, but were not rigorously monitored. In summary, on Day 1, participants were admitted to the National Institutes of Health Clinical Center after an overnight 10-12 hour fast and underwent a frequently sampled 75-gram two-hour oral glucose tolerance test (OGTT) with samples at 0, 15, 30, 60, 90, and 120 minutes to measure plasma glucose and insulin concentrations. Blood samples for genome-wide genotyping were collected once at time 0 during the first visit. On Day 2, stable isotope kinetic tracers of glucose and deuterated water were administered to measure post-absorptive rates of glucose appearance and gluconeogenesis, as previously described [5]. Serial pharmacokinetic sampling was performed after 3 months on Day 1 at 0h, 0.5h, 1h, 1.5h, 2h, 3h, 4h, 6h, 8h, 12h (pre-PM dose), 24h, and 30h post-AM dose and processed for isolation of plasma and red blood cells (Supplemental Figure 2b).

### 2.4 Analytical assays

Metformin concentrations from human plasma and red blood cell (RBC) samples were quantified using a validated HPLC-MS/MS assay over the concentration range of 10 ng/mL to 10,000 ng/mL on a Sciex QTRAP® 6500 mass spectrometer. This assay was performed at the Clinical Pharmacology Laboratory, National Institutes of Health, Clinical Center (Bethesda, MD).

Liraglutide was measured by µHPLC-MS/MS assay over the concentration range of 1– 250 ng/mL on a Sciex QTRAP® 6500 mass spectrometer. Glucose concentrations were measured in plasma using an enzymatic hexokinase assay on the Cobas 6000 instrument (Roche Diagnostics, USA). Insulin and C-peptide were measured in serum via electrochemiluminescence on the Cobas 6000 instrument (Roche Diagnostics). HbA1c was determined using the high-performance liquid chromatography (HPLC) D10 instrument (Bio-Rad, USA). Renal function was calculated using the 2009 Creatinine-based CKiD “Bedside” Equation and CKiD U25 Creatinine Equation [29].

### 2.5 Noncompartmental analysis

A non-compartmental approach to PK model analysis was employed using Phoenix WinNonlin v8.3 (Certara Corp, Cary, NC). The maximum plasma concentration (CMAX) and the time of maximum plasma concentration (TMAX) were recorded as observed values. The area under the concentration-time curve (AUC) from time zero to the time of the final quantifiable sample (AUCLAST) and from time zero to the dosing interval (AUCTAU) was calculated using the linear-up/log-down trapezoidal method (model type Plasma [200–202]). The AUC from time zero to infinity (AUCINF) was calculated by extrapolation by dividing the last measurable drug concentration (CLAST) by the rate constant of the terminal phase, λZ. This constant was determined from the slope of the terminal phase of the concentration-time curve using uniformly weighted least-squares as the estimation procedure and acceptance criteria of i) adjusted r^2^ > 0.8, ii) inclusion of > 3 time points in the terminal phase. Estimated PK parameters included the apparent volume of distribution at steady state (Vd/F) and the apparent oral systemic clearance at steady state (CL/F), which was calculated as absolute dose divided by AUCINF. The extrapolated AUCINF was >25% and the clearance and volume estimates for those subjects were flagged and excluded from statistical summaries.

### 2.6 Population pharmacokinetics analysis

Population PK model analysis and data visualization were performed using the pharmacometrics package Pumas version 2.5.1 (PumasAI, Dover, Delaware) in Julia 1.9.3 (JuliaHub, Cambridge, Massachusetts). Data processing and statistical analyses were also performed using R version 4.3.3 (R Foundation, Vienna, Austria). First-order conditional estimation methods were utilized during the model-building process. The base PK model was defined as the structural model that best described the metformin concentration-time data without consideration of covariate effects. One- and two-compartment models with zero- or first-order absorption were evaluated. Additive, proportional, and combination additive/proportional residual error models were explored to account for the intraindividual variability. Between-subject variability (BSV) using an exponential error model was evaluated on all PK parameters, assuming a log-normal distribution. Model evaluation and selection were guided by changes in the Akaike information criterion (AIC) and Bayesian information criterion (BIC), along with visual assessment of goodness-of-fit (GOF) plots (Figure 3a-d).

The covariates considered for inclusion were weight, body mass index (BMI), sex, serum creatinine, treatment group, multi-locus genetic risk score for metformin response, and individual genetic variants evaluated in the pharmacogenetic analysis. Continuous variables were explored using median-normalized power functions, with the exception of weight, which was evaluated using allometric scaling. Categorical variables were explored as proportional relationships.

Covariate selection employed stepwise forward selection and backwards elimination (*P*<0.05; delta OFV drop of 3.84 for 1 degree of freedom). Given the exploratory nature of this study and limited sample size, a *P*-value of 0.05 was deemed acceptable for identifying covariates of interest. The aim of this PK analysis was to generate hypotheses and highlight relationships that merit further research; therefore, it was essential to evaluate biologically plausible covariates that may not have met stricter criteria. Additional covariate selection criteria included reduction of BSV, and visual inspection of GOF plots (Figure 3a-d).

Reliability of the final population PK model was assessed through internal validation using a nonparametric bootstrap method. A total of 1,000 bootstrap replicates were generated from the original dataset, and the resulting median values, standard errors, and 95% confidence intervals were compared with the parameter estimates obtained from the final model. Predictive performance was further evaluated using a prediction-corrected visual predictive check (Figure 2).

**Figure 2:**
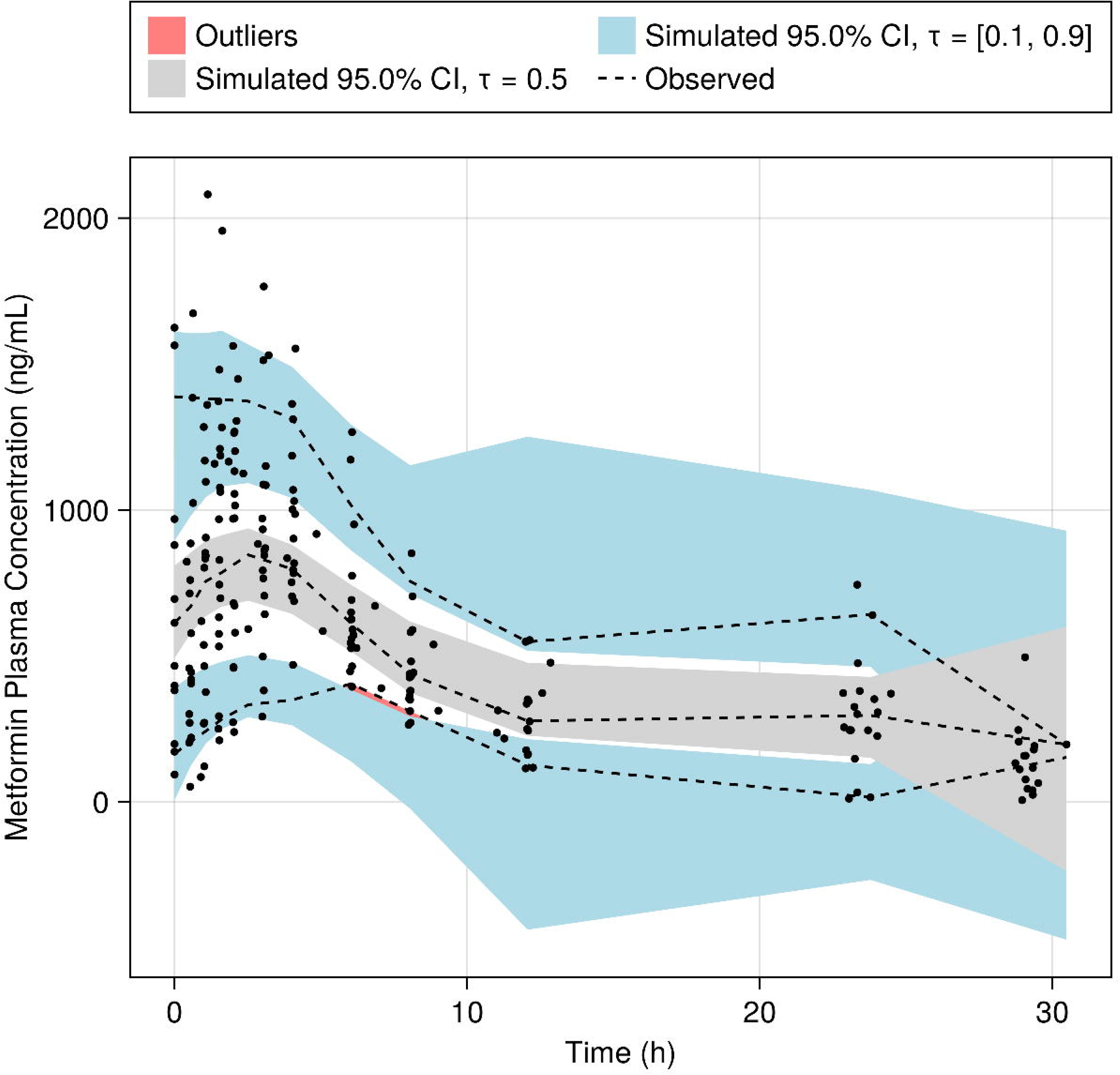
Prediction-corrected visual predictive check of metformin plasma concentrations versus time based on the final population pharmacokinetic model. The lines represent the observed 10^th^, 50^th^, and 90^th^ percentiles, and the dots denote individual observed concentration measurements. The blue shaded areas depict the simulation-based 95% confidence intervals for the 10^th^ and 90^th^ percentiles, while the gray shading represents the 95% confidence interval for the simulated 50^th^ percentile.

**Figure 3.**
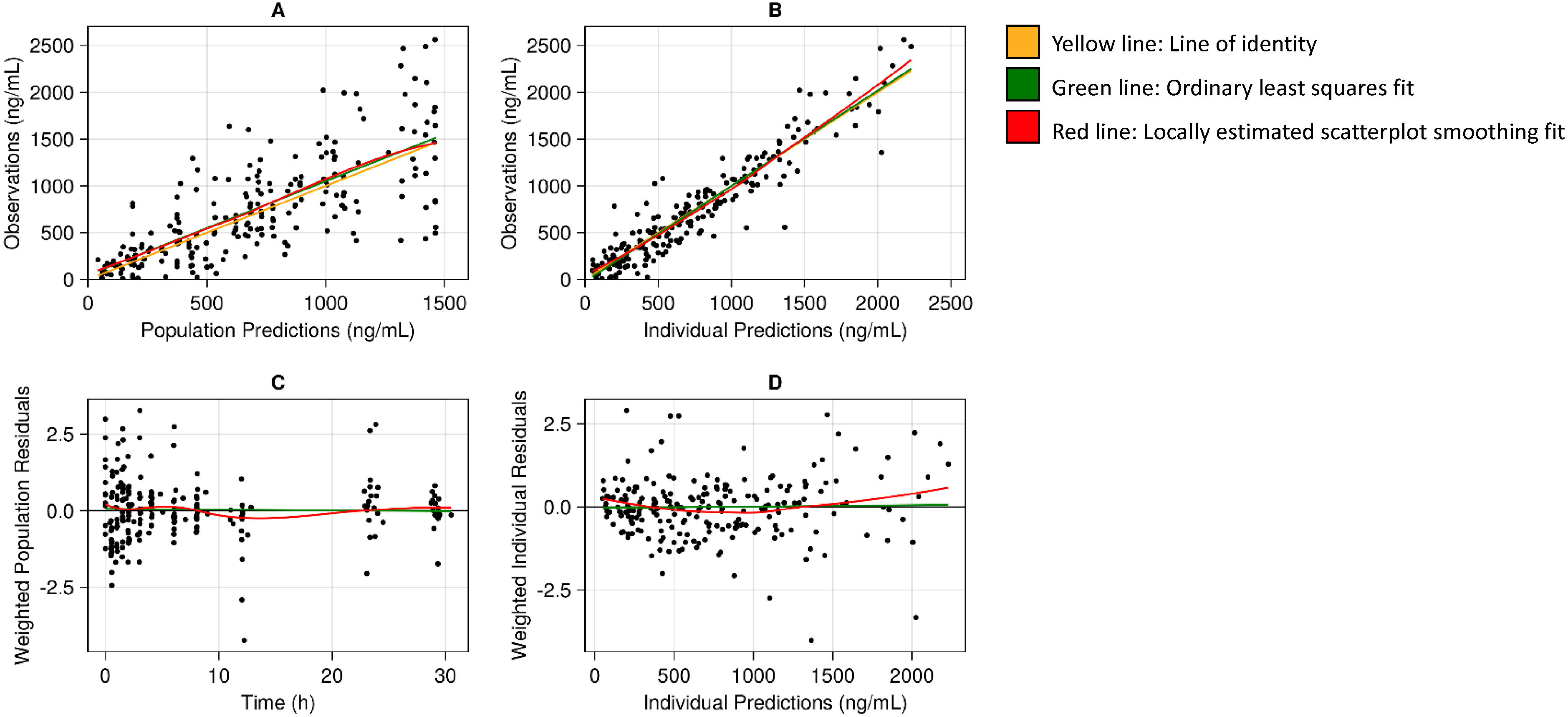
(a-d): Goodness-of-fit plots of the final metformin pharmacokinetic model. These plots illustrate the goodness-of-fit and model performance of the final model. Panel (a) displays observed concentrations vs population predicted concentrations, (b) observed concentrations vs individual predicted concentrations, (c) weighted residuals vs time, and (d) weighted individual residuals vs individual predictions. Yellow line = line of identity; Green line = OLS (ordinary least squares fit); Red line = LOESS (locally estimated scatterplot smoothing fit). Abbreviations: OLS, ordinary least squares fit; LOESS, locally estimated scatterplot smoothing fit

As some participants received asymmetric dosing (500mg in the morning and 1000mg in the evening), metformin AUC0-24 was selected over AUC0-12 to better characterize the total daily metformin exposure. The model-simulated AUC0-24 was then compared with pharmacodynamic outcomes, including changes in HbA1c, absolute rates of gluconeogenesis, and the results of the OGTT from baseline.

### 2.7 Pharmacogenetic analysis and multi-locus genetic risk score for metformin response

Genomic DNA was extracted from peripheral blood samples collected at the NIH Clinical Center. Genotyping was conducted at the NHGRI’s Genomics Core (National Human Genome Research Institute, NIH, Bethesda, MD, USA) using either Illumina’s Multi-Ethnic Genotyping Array or the Global Diversity Array, following manufacturers’ standard protocols. Samples were stored at -80° C under standard biobanking conditions prior to genotyping. Quality control was applied within and across genotyping batches, including filtering of samples based on call rate, Hardy-Weinberg Equilibrium, and concordance checks. Genotype data generated from different arrays were harmonized to a common reference build and allele coding prior to analysis to ensure comparability across platforms. Genotypes for most variants were available from the arrays (*OCT1*: rs622342, rs628031, rs461473; *OCT2*: rs316019; *MATE2*: rs12943590; *GLP-1R* (Glucagon-like peptide-1 receptor): rs6923761; *TCF7L2* (Transcription factor 7-like 2): rs7903146; *CTRB1/2* (Chymotrypsinogen B 1/2): rs7202877). Other variants were imputed using the TOPMED mixed populations reference data [30]. Imputed variants were all of high quality (INFO score >0.98).

Variants were selected for analysis based on evidence for an interaction between the variant and treatment response or complications (Table 1). These variants were further limited to those with the strongest association with metformin and liraglutide response, as evidenced from multiple studies or by genes with strong biological plausibility. In this targeted genetics approach, we investigated well-established variants in candidate genes with a minor allele frequency of >20% (*OCT1/2, GLUT2, MATE 1/2, ATM*) to evaluate whether these known variants were related to glycemic traits after metformin treatment in Y-T2D.

**Table 1:**
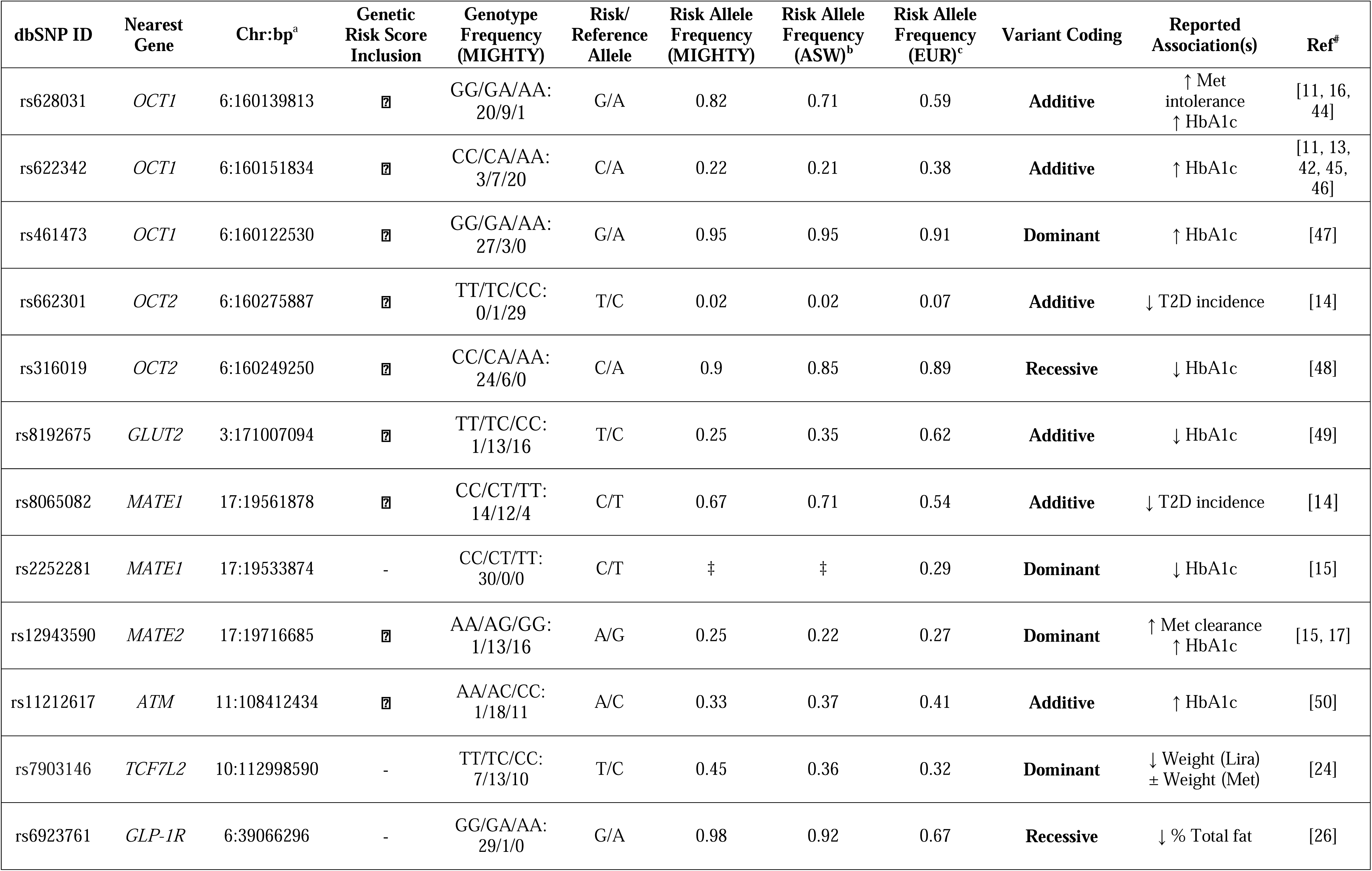
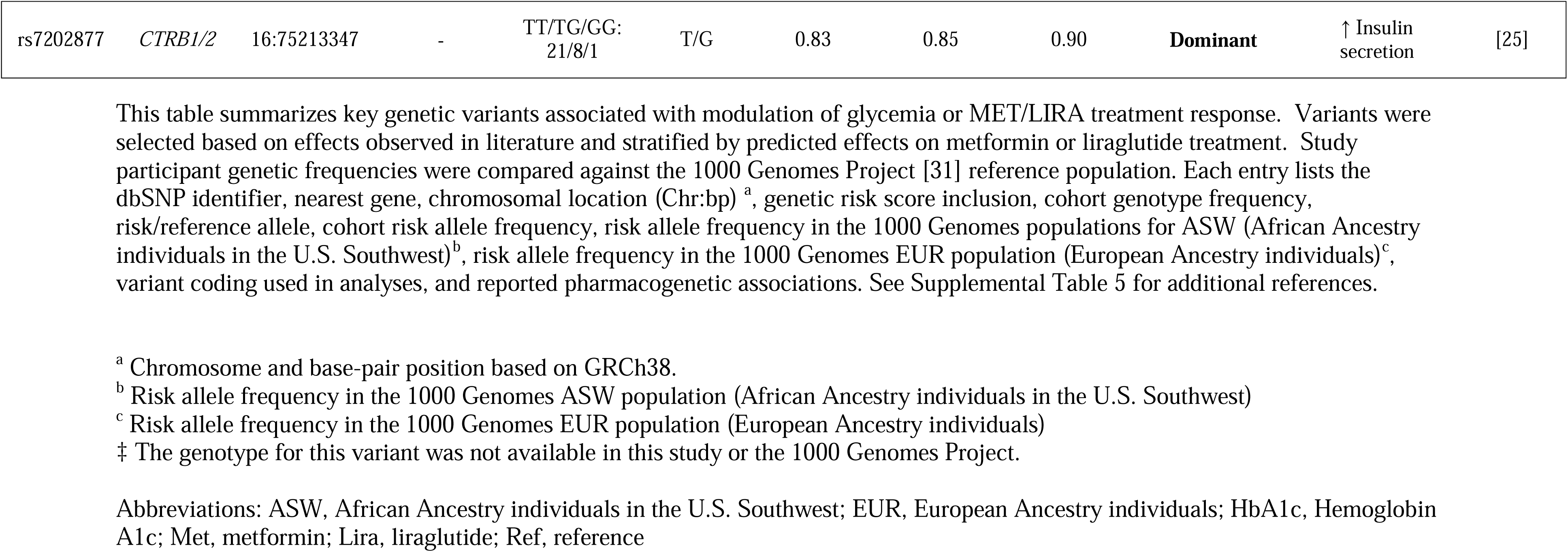
Summary of variants evaluated for pharmacogenetic effects.

| dbSNP ID | Nearest Gene | Chr:bp<sup>a</sup> | Genetic Risk Score Inclusion | Genotype Frequency (MIGHTY) | Risk/Reference Allele | Risk Allele Frequency (MIGHTY) | Risk Allele Frequency (ASW)<sup>b</sup> | Risk Allele Frequency (EUR)<sup>c</sup> | Variant Coding | Reported Association(s) | Ref<sup>#</sup> |
| --- | --- | --- | --- | --- | --- | --- | --- | --- | --- | --- | --- |
| rs628031 | *OCT1* | 6:160139813 | ☑ | GG/GA/AA:<br>20/9/1 | G/A | 0.82 | 0.71 | 0.59 | **Additive** | ↑ Met intolerance<br>↑ HbA1c | [11, 16, 44] |
| rs622342 | *OCT1* | 6:160151834 | ☑ | CC/CA/AA:<br>3/7/20 | C/A | 0.22 | 0.21 | 0.38 | **Additive** | ↑ HbA1c | [11, 13, 42, 45, 46] |
| rs461473 | *OCT1* | 6:160122530 | ☑ | GG/GA/AA:<br>27/3/0 | G/A | 0.95 | 0.95 | 0.91 | **Dominant** | ↑ HbA1c | [47] |
| rs662301 | *OCT2* | 6:160275887 | ☑ | TT/TC/CC:<br>0/1/29 | T/C | 0.02 | 0.02 | 0.07 | **Additive** | ↓ T2D incidence | [14] |
| rs316019 | *OCT2* | 6:160249250 | ☑ | CC/CA/AA:<br>24/6/0 | C/A | 0.9 | 0.85 | 0.89 | **Recessive** | ↓ HbA1c | [48] |
| rs8192675 | *GLUT2* | 3:171007094 | ☑ | TT/TC/CC:<br>1/13/16 | T/C | 0.25 | 0.35 | 0.62 | **Additive** | ↓ HbA1c | [49] |
| rs8065082 | *MATE1* | 17:19561878 | ☑ | CC/CT/TT:<br>14/12/4 | C/T | 0.67 | 0.71 | 0.54 | **Additive** | ↓ T2D incidence | [14] |
| rs2252281 | *MATE1* | 17:19533874 | - | CC/CT/TT:<br>30/0/0 | C/T | ‡ | ‡ | 0.29 | **Dominant** | ↓ HbA1c | [15] |
| rs12943590 | *MATE2* | 17:19716685 | ☑ | AA/AG/GG:<br>1/13/16 | A/G | 0.25 | 0.22 | 0.27 | **Dominant** | ↑ Met clearance<br>↑ HbA1c | [15, 17] |
| rs11212617 | *ATM* | 11:108412434 | ☑ | AA/AC/CC:<br>1/18/11 | A/C | 0.33 | 0.37 | 0.41 | **Additive** | ↑ HbA1c | [50] |
| rs7903146 | *TCF7L2* | 10:112998590 | - | TT/TC/CC:<br>7/13/10 | T/C | 0.45 | 0.36 | 0.32 | **Dominant** | ↓ Weight (Lira)<br>± Weight (Met) | [24] |
| rs6923761 | *GLP-1R* | 6:39066296 | - | GG/GA/AA:<br>29/1/0 | G/A | 0.98 | 0.92 | 0.67 | **Recessive** | ↓ % Total fat | [26] |
| rs7202877 | *CTRB1/2* | 16:75213347 | - | TT/TG/GG:<br>21/8/1 | T/G | 0.83 | 0.85 | 0.90 | **Dominant** | ↑ Insulin<br>secretion | [25] |

<sup>a</sup> Chromosome and base-pair position based on GRCh38. <sup>b</sup> Risk allele frequency in the 1000 Genomes ASW population (African Ancestry individuals in the U.S. Southwest) <sup>c</sup> Risk allele frequency in the 1000 Genomes EUR population (European Ancestry individuals) ‡ The genotype for this variant was not available in this study or the 1000 Genomes Project.
Abbreviations: ASW, African Ancestry individuals in the U.S. Southwest; EUR, European Ancestry individuals; HbA1c, Hemoglobin A1c; Met, metformin; Lira, liraglutide; Ref, reference

Multi-locus genetic risk scores for metformin response were constructed based on variants previously reported to be associated with metformin response or tolerance (Table 1). Variants were coded in terms of genotype or allele associated with increased risk (e.g., higher HbA1c or T2D), even if that was the allele with higher frequency. As variants were all selected to have large effects, all were weighted equally. The genetic risk score for each individual was the sum of risk alleles across all variants, in line with the hypothesis that the score would be associated with outcomes in the direction corresponding to higher glycemia and diabetes risk. Details regarding variants, allele coding, risk allele, and risk allele frequency in this population and in reference populations are given in Table 1. All individuals had the genotype associated with increased risk for rs461473; therefore, this variant was excluded from the genetic risk score calculation.

### 2.8 Statistical analysis

This was a pre-specified exploratory analysis, and sample size calculations were based on the study’s previously reported primary outcome difference in gluconeogenesis between MET and MET+LIRA [5]. Data are presented as mean (95% Confidence Interval) unless otherwise stated. No data were imputed, and only per-protocol analysis was performed. Glycemic outcomes (dependent variables) were fasting glucose and rates of fasting gluconeogenesis. Independent (predictor) variables were pharmacokinetic (Ka, CL/F, Vd/F) and pharmacogenomic (genetic risk score, individual genetic variants). Ordinary least squares multilinear regression models were created to evaluate predictors of fasting glucose and fractional gluconeogenesis. A baseline model with the biological predictors age, biological sex, treatment group, and baseline value (Model 1) was created first to establish a baseline. Subsequent models included genetic risk score (Model 2), rs628031_*OCT1* (Model 3), and rs622342_*OCT1* (Model 4) with covariates age, biological sex, treatment group, and baseline value. To evaluate whether genetic variants were serving as a proxy for genome-wide genetic ancestry (e.g. proportion genome-wide African ancestry), sensitivity analyses were conducted in which models were adjusted for the first two principal components of the genotypes. Statistical significance was defined as *P*-value <0.01, adjusting for multiple comparisons with Bonferroni correction. Statistical analyses were performed with STATA, v18.0 (College Station, Texas).

## 3. Results

The baseline participant characteristics are summarized in Table 2, with no difference in characteristics between the genetics cohort and the subgroup with PK data (Supplemental Tables 2 and 3). In the participants who completed the randomized trial, 80% were on MET 1500-2000 mg/day or LIRA >1.2mg daily. Minor gastrointestinal side effects (nausea, abdominal pain, diarrhea) occurred in 55% (12/22).

**Table 2:**
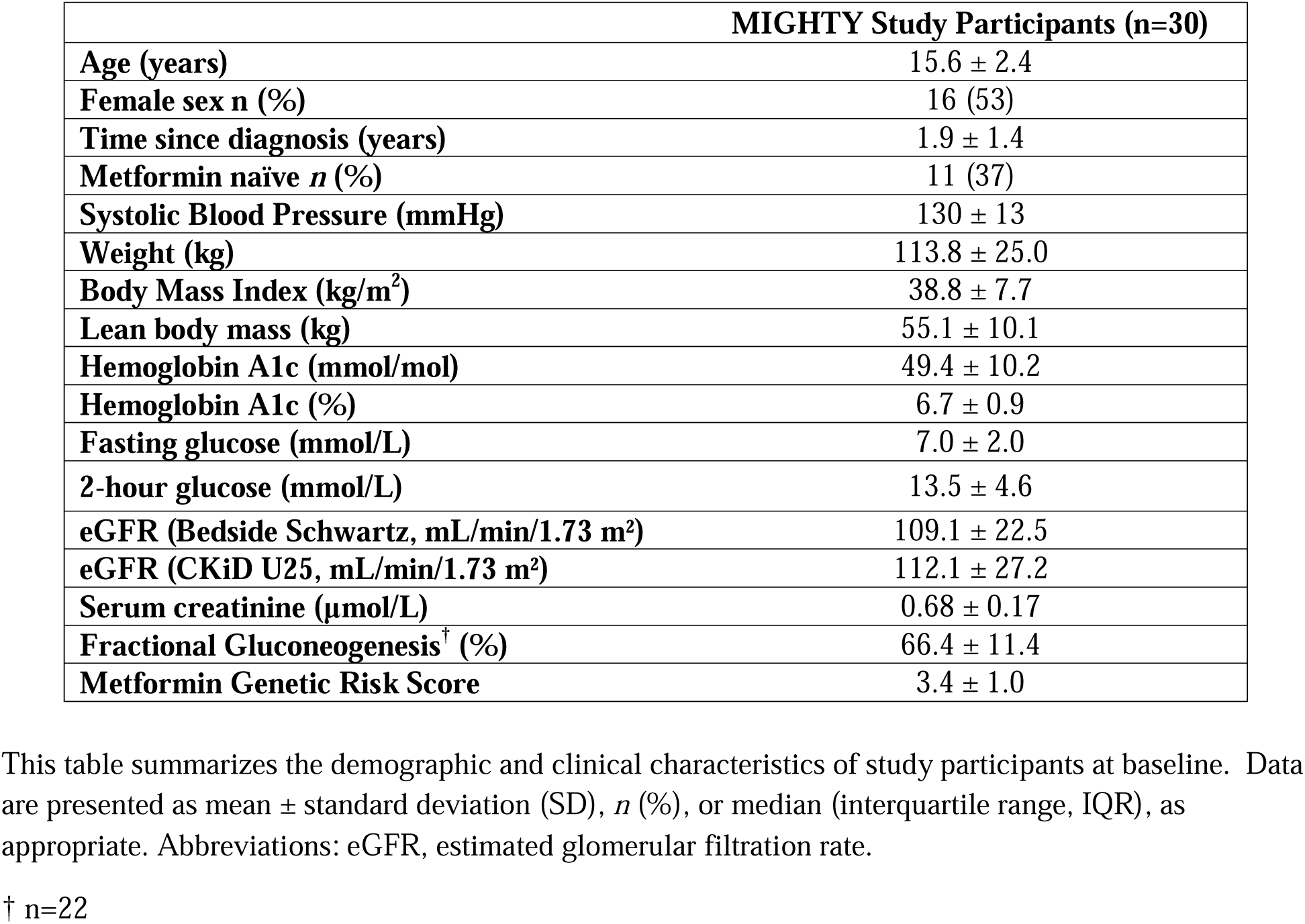
Baseline Participant Characteristics.

**Table 3:** Pharmacokinetic model parameters.

| Parameter (unit) | Estimate | SE | Bootstrap median (95 % CI) |
| --- | --- | --- | --- |
| **Fixed Effects** |  |  |  |
| **K<sub>a</sub>: Absorption rate constant (hr<sup>-1</sup>)** | 0.19 | 0.02 | 0.20 (0.16-0.26) |
| **CL/F: Apparent oral clearance (L/hr)** | 86.69 | 8.47 | 87.00 (71.43-105.13) |
| **Effect of *OCT1*_rs628031 on CL/F** | 0.58 | 0.35 | 0.55 (0.17-1.34) |
| **Vd/F: Apparent oral volume of distribution (L)** | 170.25 | 47.96 | 173.47 (102.52-295.61) |
| **Between-subject variability** |  |  |  |
| $\omega^2_{K_a}$ | 0.09 | 0.06 | 0.06 (0.00-0.20) |
| $\omega^2_{CL}$ | 0.10 | 0.03 | 0.09 (0.04-0.15) |
| $\omega^2_{Vd}$ | 0.77 | 0.32 | 0.67 (0.18-1.44) |
| **Residual error** |  |  |  |
| $\sigma^2_{Additive}$ | 40308 | 9234.7 | 40078.5 (25428-60428) |

This table summarizes the estimated pharmacokinetic parameters from the constructed population pharmacokinetic model. Values are presented as the population mean estimate, standard error (SE), and corresponding 95% confidence interval (CI). $\omega^2$ represents the variance of each parameter. Abbreviations: K<sub>a</sub>, absorption rate constant; CL, clearance; Vd/F, volume of distribution; $\omega^2$ , interindividual variability; $\sigma^2$ , intraindividual variability SE, standard error; CI, confidence interval

The frequencies of pre-selected variants in our cohort were comparable to those of African Americans enrolled in the 1000 Genomes Project [31] (Table 1). Principal component analysis of the genome-wide genotypes indicates that study samples cluster along a continuum between African ancestry individuals and European ancestry individuals, as expected of individuals who self-identify as African American (Figure 4).

**Figure 4:**
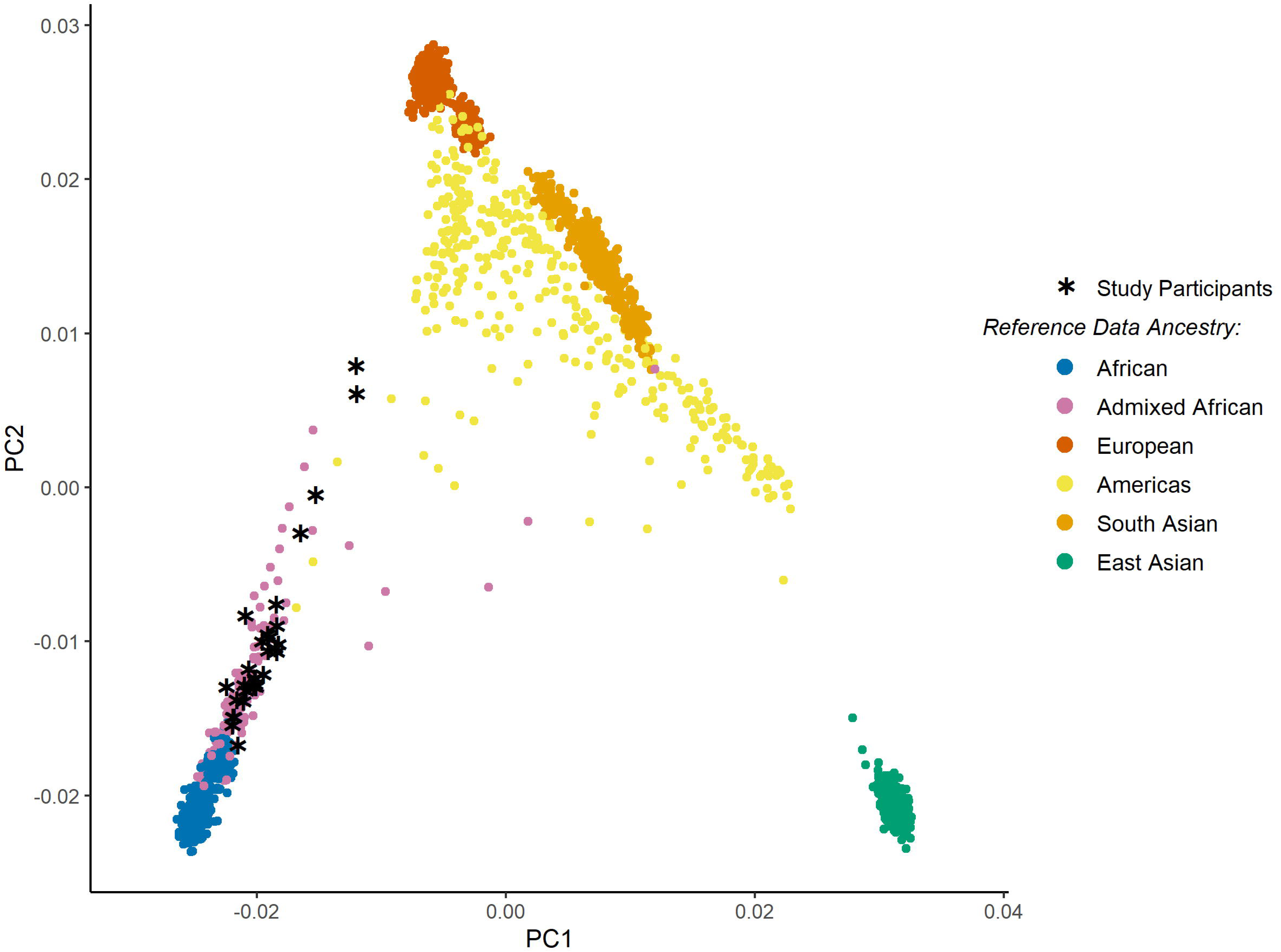
Principal Component Analysis of Study Participants with 1000 Genomes Reference Data. Shown are the first 2 principal components of the genotypes for study participants (n=30) as well as participants of the 1000 Genomes Data (The 1000 Genomes Project Consortium. A global reference for human genetic variation. *Nature* **526**, 68–74 (2015). https://doi.org/10.1038/nature15393). 1000 Genomes Data are colored according to genetic ancestry by super-populations (as described in [same citation]), except that admixed African ancestry individuals (African Americans and African Caribbeans) are identified separately from African ancestry for comparison to study population. The study participants (denoted by asterisks) are distributed along the axis between African and European ancestry clusters, overlapping the admixed African ancestry and indicating capture of the genetic ancestry variation in the African American population.

### 3.1 One-compartment metformin PK model

Plasma concentrations of metformin were best described by a one-compartment model with first-order absorption and elimination, along with an additive residual error model. Between subject variability was included on Ka, Vd/F, and CL/F. Pharmacokinetic parameters stratified by treatment group are shown in Supplemental Table 1. While metformin concentrations were also measured in RBCs as a potential deep compartment, the values were highly variable (Supplemental Figure 3), and incorporating this compartment into the model resulted in reduced model stability, physiologically implausible parameter estimates, and poor predictive performance. Therefore, model development focused on plasma data.

In the covariate analysis, effects of the rs628031_*OCT1* variant were included as a significant covariate on CL/F (*P* = 0.01; ΔOFV = 6.52). Final PK model parameter estimates and results of the bootstrap are shown in Table 3 and GOF plots can be found in Figure 3a-d. Modeling was also conducted without pharmacogenetic variants, and the unadjusted CL/F was 103.2 L/hr. A small subset of participants met hyperfiltration criteria (≥140 mL/min/1.73 m²) at baseline and post-treatment; however, eGFR was not associated with clearance (*P*>0.05, data not shown). Parameter estimates from the final PK model were within the 95% confidence interval derived from the bootstrap replicates, confirming model robustness. Additionally, the prediction-corrected visual predictive check demonstrated that the observed concentrations were well contained within the simulated 90% prediction intervals as shown in Figure 2. Ka, Vd/F, CL/F, and metformin AUC did not correlate with glycemia (HbA1c, gluconeogenesis, or glucose AUC during OGTT) (*P*>0.05, data not shown).

### 3.2 Determinants of fasting glucose and gluconeogenesis

Table 4 depicts the relationships between biological and genetic variables and glycemic markers. Baseline fasting glucose was significantly associated with post-treatment fasting glucose, accounting for ∼60% of variance (all *P<0.001*). Baseline fractional gluconeogenesis was associated with fractional gluconeogenesis after treatment (all *P<0.001*), as was rs622342_*OCT1* (*OCT1*) as shown in Model 4, Table 4. Individuals with the C allele for rs622342_*OCT1* had ∼8% lower rates of gluconeogenesis after treatment with MET or MET+LIRA, even after adjusting for baseline rates (*P=0.003*). Fifty-six percent (56%) of the variability in post-treatment fractional gluconeogenesis was explained by rs622342 and covariates: baseline gluconeogenesis, age, sex, and treatment group. Other individual genetic metformin variants, GRS, and PK model parameters were not associated with baseline or post-treatment fasting glucose or gluconeogenesis.

**Table 4:** Biological predictors of glycemia in youth treated with either MET or MET+LIRA.

| Model Metrics | Post-Treatment<br>Fasting Glucose (mmol/L)<br>N=24 |  | Post-Treatment<br>Fractional Gluconeogenesis (%)<br>N=21 |  |
| --- | --- | --- | --- | --- |
|  | β (95% CI) | P-value | β (95% CI) | P-value |
| MODEL 1 |  |  |  |  |
| Baseline value | 1.13 (0.71, 1.55) | <0.0001 | 0.49 (0.04 - 0.95) | 0.035 |
| Treatment group | -1.46 (-3.25, 0.34) | 0.106 | 0.10 (-9.60 – 9.39) | 0.982 |
| Age | -0.15 (-0.57, 0.27) | 0.455 | -1.12 (-3.39 – 1.15) | 0.311 |
| Sex | -1.88 (-3.61, -0.15) | 0.035 | -4.83 (-15.77 – 6.11) | 0.363 |
|  | Adjusted R<sup>2</sup>=0.5757, P-value= 0.0003 |  | Adjusted R<sup>2</sup>=0.2476, P-value= 0.072 |  |
| MODEL 2 |  |  |  |  |
| Genetic Risk Score | 0.79 (-0.08, 1.66) | 0.073 | -3.02 (7.68 – 1.64) | 0.188 |
| Baseline value | 1.22 (0.81, 1.63) | <0.0001 | 0.49 (0.05 – 0.93) | 0.032 |
| Treatment group | -1.10 (-2.84, 0.63) | 0.198 | -1.65 (-11.24 – 7.93) | 0.719 |
| Age | -0.23 (-0.64, 0.17) | 0.243 | -0.83 (-3.10 – 1.43) | 0.446 |
| Sex | -2.01 (-3.64, -0.37) | 0.019 | -4.11 (-14.87 – 6.64) | 0.438 |
|  | Adjusted R<sup>2</sup>=0.6272, P-value= 0.0002 |  | Adjusted R<sup>2</sup>=0.2879, P-value= 0.0680 |  |
| MODEL 3 |  |  |  |  |
| rs628031_OCT1 | 1.24 (-0.46, 2.94) | 0.143 | -5.68 (-14.31 – 2.96) | 0.181 |
| Baseline value | 1.22 (0.79, 1.65) | <0.0001 | 0.49 (0.05 – 0.93) | 0.032 |
| Treatment group | -0.98 (-2.84, 0.88) | 0.285 | -2.48 (-12.43 – 7.47) | 0.603 |
| Age | -0.19 (-0.60, 0.23) | 0.355 | -1.01 (-3.23 – 1.21) | 0.346 |
| Sex | -2.02 (-3.71, -0.34) | 0.021 | -4.28 (-15.00 – 6.43) | 0.408 |
|  | Adjusted R<sup>2</sup>=0.6036, P-value= 0.0004 |  | Adjusted R<sup>2</sup>=0.2904, P-value= 0.0666 |  |
| MODEL 4 |  |  |  |  |
| rs622342_OCT1 | -0.05 (-1.33, 1.24) | 0.940 | -8.80 (-14.13 – -3.47) | 0.003 |
| Baseline value | 1.13 (0.69, 1.56) | <0.0001 | 0.53 (0.18 – 0.88) | 0.006 |
| Treatment group | -1.48 (-3.40, 0.45) | 0.124 | -4.79 (-12.62 – 3.04) | 0.212 |
| Age | -0.15 (-0.59, 0.28) | 0.466 | -1.12 (-2.86 – 0.63) | 0.192 |
| Sex | -1.87 (-3.66, -0.08) | 0.041 | -3.53 (-11.98 – 4.92) | 0.387 |
|  | Adjusted R<sup>2</sup>=0.5522, P-value= 0.0011 |  | Adjusted R<sup>2</sup>=0.5601, P-value= 0.0028 |  |

This table summarizes multilinear regression models evaluating biological predictors of fasting glucose and fractional gluconeogenesis (GNG) in youth treated with metformin and with or without liraglutide. Each model reports the estimated $\beta$ coefficient (95% confidence interval) and corresponding P-value for each predictor. Adjusted R<sup>2</sup> and overall model P-values are shown below each model. Models include the covariates baseline glycemic value, treatment group (MET vs. MET+LIRA), age, and sex. Model 1 assessed the influence of baseline glycemia. Model 2 includes a multi-locus genetic risk score for metformin response and showed associations with fasting glucose and fractional gluconeogenesis. These associations were delineated in Models 3 and 4, which used the OCT1 variants rs628031 and rs622342. Abbreviations: GNG, gluconeogenesis; $\beta$ , regression coefficient; CI, confidence interval; OCT1, organic cation transporter 1

We also found a nuanced association with the T allele of rs7903146_*TCF7L2* and lower fasting glucose. When considering both the MET and MET+LIRA treatment groups together, rs7903146_*TCF7L2* was not significantly associated with fasting glucose, but there was effect modification by group (*P*=0.06, Supplemental Table 4). In a sensitivity analysis in youth treated with MET+LIRA only, the T allele for rs7903146_*TCF7L2* tended to have a lower fasting glucose ((β = -1.32 [-2.42, -0.22] mmol/L, Adjusted R^2^ = 0.8, *P* < 0.002) Supplemental Table 4). No other variants related to liraglutide response were significantly associated with baseline or post treatment fasting glucose or gluconeogenesis (*P*>0.05, data not shown). All effect estimates of genetic associations were unchanged in sensitivity analyses adjusting models for genome-wide genetic ancestry (the first two principal components of the genotypes), indicating that the genetic associations observed were not a proxy for genetic ancestry.

## 4. Discussion

This study investigated pharmacokinetic and pharmacogenetic predictors of fasting glycemic traits after short-term metformin and liraglutide in a well characterized cohort of African American Y-T2D. A one-compartment population PK model with first-order absorption and elimination, additive residual error, and rs628031_*OCT1* as a covariate, best described metformin pharmacokinetics in African American Y-T2D. Notably, the C allele of rs622342_*OCT1* was associated with an 8% reduction in fractional gluconeogenesis independent of baseline glycemia, indicating genetic variation in metformin’s hepatic mechanism of action.

These findings highlight the power of utilizing a population PK model in Y-T2D to integrate metformin PK parameters with demographic and genetic variables. Contrary to our hypothesis, neither metformin clearance nor genetic risk score predicted changes in fasting plasma glucose following 3 months of treatment. This discrepancy is potentially explained by highly conserved glycemic pathways. Pathway redundancy is common in biological systems. Although the structural *OCT1* variant rs628031 was shown to modulate both hepatic transport and glycemic outcomes in murine models [20, 21], in humans, multiple transporters are implicated in metformin transport [32] and metformin clearance may be decoupled from uniform glycemic changes in Y-T2D. Additionally, our genetic risk score was constructed with a priori selection of variants that were implicated in altered adult metformin response which may not mirror causation for metformin failure in pediatric and adolescent populations. The pathophysiology of Y-T2D has an established profile that contrasts from type 2 diabetes in adults [3, 4]; therefore, our findings may reflect a critical direction for further studies of metformin treatment failure. Interestingly, in the relatively small group treated with a combination of metformin and liraglutide, we found an association between a genetic variant in *TCF7L2* and reductions in fasting glucose in Y-T2D, an observation that requires further exploration. Baseline glycemia remained the strongest predictor of treatment response, a finding that aligns with large Y-T2D multi-center studies [3, 33]. These nuanced associations with two established *OCT1* variants suggest that genetic variability is important for pharmacokinetic and mechanistic features of metformin response in Y-T2D.

This study adds three new contributions to improve our understanding of treatment responsiveness in Y-T2D. First, genetic variation was associated with differences in rates of gluconeogenesis among metformin treated Y-T2D. Excessively high rates of gluconeogenesis are a primary pathological characteristic of Y-T2D [5, 27], and this novel finding emphasizes the role of *OCT1* genetic variants as possible mediators of metformin’s ability to lower gluconeogenesis in Y-T2D. Finding an association between gluconeogenic flux and rs622342*_OCT1* provides additional evidence to corroborate previous studies that linked rs622342_*OCT1* C allele carriers with reduced hepatic insulin resistance, improved glycemic control (fasting glucose, HbA1c), and higher metformin plasma concentrations [17, 34–36]. Our findings provide mechanistic insight, albeit over 3 months, to support recent analyses in a large longitudinal clinical trial of 1285 patients enrolled in the Rotterdam Study for 27 years, that showed improved HbA1c response in rs622342 C allele carriers [36]. We posit that the observed short-term reductions in gluconeogenesis after three months may translate to long-term glucose reductions. These findings contrast with earlier reports that observed higher HbA1c or no effect of the C allele on metformin response, (Table 1) [13, 20, 37, 38]. Inconsistent study findings may be secondary to biases in study design (small sample sizes, especially among individuals with modest elevations in HbA1c in whom large changes in HbA1c are unlikely) or biological factors (redundancy in cation transporters *in vivo* with compensation to preserve functional effects). Although *in vitro* hepatocyte cell lines and *in vivo* mouse models implicate *OCT1* as critical for hepatic uptake of metformin and lowering of glucose production [20], differential effects of metformin on gluconeogenesis in humans may depend on the fasting duration and the presence or absence of diabetes. For example, after a prolonged 48-hour fast in healthy persons, rates of gluconeogenesis were higher in metformin-treated group compared to controls and no differences in glucose production were observed by *OCT1* genotype [39]. Additionally, rs622342_*OCT1* is an intronic variant, and may be associated more closely with *OCT1* expression instead of function, potentially explaining variability across studies [32].

Second, genetic contributions to metformin pharmacokinetics and treatment response are subtle and likely influenced by both genotype and drug interactions. Metformin pharmacokinetic parameters in Y-T2D were comparable to the few published estimates in youth with obesity and adults with Y-T2D [19, 40]. The metformin clearance rate was at the reported upper limit of population ranges (103.2 L/hr vs ranges 52.6 L/hr-102 L/hr reported in the literature [41]), though 20% of Y-T2D had hyperfiltration (eGFR ≥140 mL/min/1.73 m²). Greater metformin clearance (∼40%) was observed in A-allele carriers of rs628031_*OCT1*. Higher clearance among A-allele carriers could reflect either: (1) enhanced OCT1-mediated hepatic uptake (favorable for efficacy, as hepatic uptake is critical for metformin’s suppression of gluconeogenesis), or (2) increased renal elimination (unfavorable, due to reduced systemic and hepatic exposure). The A allele of the rs628031_*OCT1* variant has been associated with a lower odds of metformin-induced gastrointestinal issues [16], which may reflect the greater clearance we found, though our study was not powered to observe these effects. Clinical translation of the relationship between metformin clearance and the rs628031_*OCT1* genotype remains unclear and warrants further investigation, but our findings align with the only other pharmacokinetic-pharmacogenetic study in adolescents, which found no relationship between *rs628031_OCT1* and glycemic outcomes after 6 months of metformin treatment [19].

Our study also highlights the importance of exploring pharmacogenetic associations in Y-T2D treated with a GLP-1RA. Examining variants that may influence response to combination therapies is a critical step towards optimizing therapeutic strategies, motivating our analysis of risk variants in *GLP1R*, which encodes the GLP-1 receptor, and *TCF7L2*, a transcription factor implicated in global T2D risk, insulin secretion, GLP-1R expression, and endogenous GLP-1 production [24–26] (Table 1). Among those in the MET+LIRA treatment group, rs7903146_*TCF7L2* tended to be associated with improved fasting glucose (*P*=0.028) but failed to meet the Bonferroni-corrected significance threshold of *P*<0.01. Mechanistically, a deleterious *TCF7L2* variant could be associated with greater sensitivity to an exogenous GLP-1RA due to impairment of endogenous production pathways. Therefore, the within-group improvement for MET+LIRA treatment provides a compelling hypothesis that must be assessed in more rigorous models.

Finally, our deep phenotyping study extended prior observations that support baseline fasting glucose and gluconeogenesis as the strongest predictors of treatment response, independent of pharmacokinetic and genomic factors. Our findings align with the TODAY trial—which similarly did not replicate many of the metformin response associations observed in adults [8]. Importantly, the TODAY study did not conduct a priori genetic analyses, all study subjects were within two years of diagnosis, and a static threshold of treatment failure was defined as HbA1c >64 mmol/mol (8%) or metabolic decomposition requiring insulin. Our novel findings offer an expansion beyond their work, including examining mechanisms in Y-T2D with more advanced pathogenesis (HbA1c <75 mmol/mol (9%), and examination of relationships along a continuum of glycemia. Pharmacogenetic analysis of hepatic gluconeogenic flux, a distinctive phenotypic feature of Y-T2D, is of particular importance for advancing mechanistic insights for treatment variability. Collectively, these results suggest that genetic risk scores based on known variants in these genes may not be clinically advantageous for predicting metformin response in Y-T2D, but candidate-gene population-specific analysis could help to shed light on variations in genotype-phenotype correlation by ancestry. Notably, many studies that observed deleterious associations were conducted in individuals primarily of European ancestry, limiting their generalizability to our study. For example, both the C allele of rs622342*_OCT1* and the A allele of rs628031_*OCT1* are observed in 10-20% lower frequency among African Ancestry individuals compared to European ancestry individuals (Table 1), highlighting a possible driver of population-level differences in treatment response in youth. In contrast, more recent studies show that previously labeled ‘deleterious risk’ alleles, such as the C-allele rs622342*_OCT1*, are protective in adults of Chinese descent [35, 42] and Mexican Mestizo descent [17].

Interestingly, the one-compartment metformin pharmacokinetic model contrasted with findings from adult studies which previously demonstrated that a two-compartment model best accounted for metformin distribution into red blood cell compartments. We found no added benefit to including a second compartment representing metformin storage in RBCs due to increased model instability (Supplemental Figure 3). The one-compartment model may be preferred in youth because the rate of transport between plasma and red blood cells may be more dependent on diffusion [43] or because of unique differences in membrane permeability among Y-T2D.

Study limitations include a small sample size that limits generalizability and reduces power to detect all relevant genetic variants or to identify smaller effect sizes on the parameters of interest. Lastly, this was a single-site study, which enabled rigorous procedural control, but our results may be confounded by regional variations in genetic and microbiome composition [27].

## 5. Conclusion

Genetic variants in the *OCT1* family (rs622342, rs628031) were common in African American Y-T2D and were related to metformin clearance and glucose indices (fasting gluconeogenesis) in metformin-treated youth. Although metformin genetic risk scores and candidate-gene variants did not predict changes in fasting glucose, the *TCF7L2* variant rs7903146 was associated with lower fasting glucose in Y-T2D treated with metformin and liraglutide. Our findings also suggest a disconnect between pharmacokinetics and glycemia in youth treated with metformin, a compelling decoupling that may be age- and disease-specific. Large-cohort longitudinal characterization of genotype and drug interactions is needed to address metformin treatment failure within the context of these findings and evaluate the functional effects on glycemia across long-term therapy. Thorough mechanistic investigation of the clinical response to metformin, especially in diverse populations, could offer improved insights into the effects of metformin and incretin-based therapies.

## Supporting information

Graphical abstract

## Acknowledgements

Thank you to Annieka Reno for her help with pharmacokinetic modeling in this project. A special thank you to the volunteers and their families whose participation made this study possible. We thank the nurses and staff at the NIH Clinical Center Metabolic Research Unit for their invaluable contributions to study procedures and protocol implementation.

## Data Availability Statement

Some or all datasets generated during and/or analyzed during the current study are available from the corresponding author on reasonable request. Genetic data is in the process of being deposited in dbGaP using controlled access, consistent with the consent form. The dbGaP accession information will be provided prior to publication.

## Authorship contribution statement

The study was conceived by CR, AC, AB and SChung. Funding was obtained and the study was administered by SChung. The study was designed by CR, WD, AC, AB and SChung, with methodological contributions from YZ, OA, WF, HC, MW, PW, ES and AB. Data acquisition, curation and resources were provided by SC, YZ, OA, WF, FD, SG, NM, IK, LM, NMal, AA, CR and SChung. HC, MW, PW and ES also contributed to data acquisition. Formal analysis was performed by SC, YZ, OA, WF, FD, SG, NM, IK, LM, NMal, AA, HC, MW, PW, ES, AB and SChung. Software and visualization were developed by SC, YZ, OA and WF, with additional visualization contributions from AB. Validation was carried out by YZ, OA, WF, HC, MW, PW, ES and AB. Investigation was led by SChung. SC, AB and SChung drafted the manuscript, and all authors critically revised it for important intellectual content. CR, WF and SChung supervised the work. All authors approved the final version of the manuscript for the integrity of the work as a whole. SChung takes responsibility for the integrity of the work and conduct of the study as a whole.

## Disclosure of Results

These data were presented during poster presentation at The 2025 Pediatric Endocrine Society Annual Meeting (Bar Harbor, MD) and oral abstract presentation at The 2025 American Diabetes Association Scientific Sessions Conference in (Chicago, IL).

## Ethics declarations

Written informed consent, and assent as applicable, were obtained prior to the start of trial-related procedures. The protocol was approved by the National Institutes of Health Institutional Review Board and registered at ClinicalTrials.gov (NCT02960659).

## Abbreviations

AIC: Akaike information criterion

ATM: Ataxia telangiectasia mutated

AUC: Area under the curve

AUC0-12: Area under the plasma concentration versus time curve from 0 to 12 hours

AUC0-24: Area under the plasma concentration versus time curve from 0 to 24 hours

AUCINF: Area under the plasma concentration versus time curve to time infinity

AUCLAST: Area under the plasma concentration versus time curve from time zero to the time of final quantifiable sample

AUCTAU: Area under the plasma concentration versus time curve during the dosing interval at steady state

BIC: Bayesian information criterion

BMI: Body mass index

BSV: Between-subject variability

CI: Confidence interval

CLAST: Last measurable drug concentration

CL/F: Apparent oral systemic clearance at steady state

CMAX: Maximum plasma concentration

CMIN: Minimum plasma concentration

CTRB1/2: Chymotrypsinogen B 1/2

eGFR: Estimated glomerular filtration rate

GLP-1: Glucagon-like peptide-1

GLP-1RA: Glucagon-like peptide-1 receptor agonist

GNG: Gluconeogenesis

GOF: Goodness of fit

GRS: Genetic risk score

HbA1c: Hemoglobin A1c

Ka: Absorption rate constant

LIRA: Liraglutide

MATE: Multidrug and toxin extrusion protein

MIGHTY: Metformin Influences Gut Hormone in Youth

MET: Metformin

NIDDK: National Institute of Diabetes and Digestive and Kidney Diseases

OCT: Organic cation transporter

OFV: Objective Function Value

OGTT: Oral Glucose Tolerance Test PK: Pharmacokinetic

PD: Pharmacodynamic

PG: Pharmacogenetic

RBC: Red blood cells

TCF7L2: Transcription Factor 7-like 2

T1/2: Terminal elimination half-life

T2D: Type 2 diabetes mellitus

TMAX: Time to maximum plasma concentration

HPLC-MS/MS: High-performance liquid chromatography with tandem mass spectrometric detection

Vd/F: Apparent oral volume of distribution

Y-T2D: Youth-onset type 2 diabetes

ω^2^: Interindividual variability (variance)

λ_Z_: Lambda Z, aka elimination rate (kEL)

