## Supplementary material for "*OCT1* Variants Are Associated with Metformin Clearance and Gluconeogenesis: Mechanistic Insights for Youth-Onset Type 2 Diabetes in the MIGHTY Study": Graphical abstract

### Slide 1
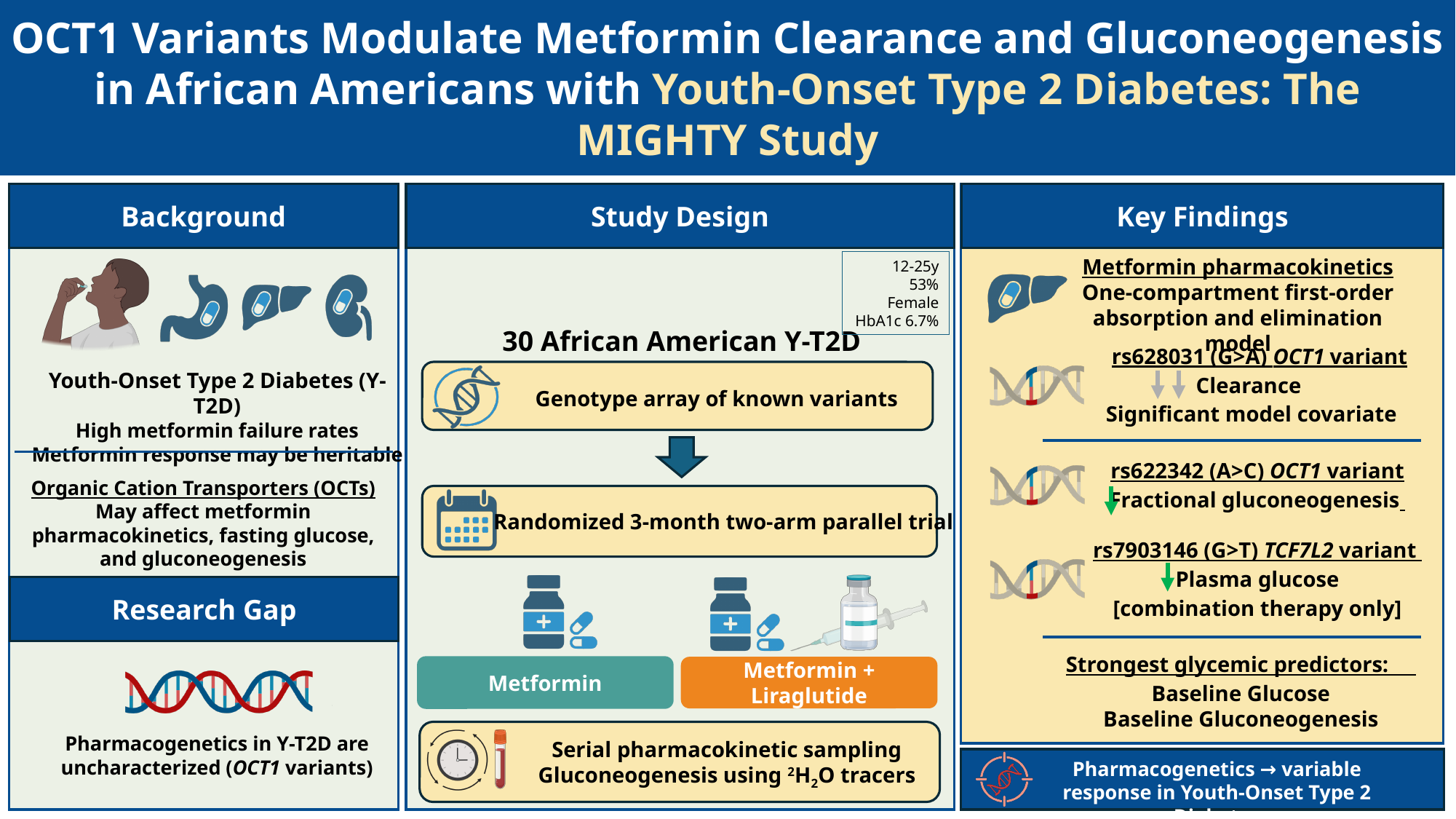

OCT1 Variants Modulate Metformin Clearance and Gluconeogenesis in African Americans with Youth-Onset Type 2 Diabetes: The MIGHTY Study
Background
Study Design
Key Findings
Metformin pharmacokinetics
One-compartment first-order absorption and elimination model
12-25y
 53% Female
HbA1c 6.7%
30 African American Y-T2D
 rs628031 (G>A) OCT1 variant
Clearance
 Significant model covariate
Genotype array of known variants
Youth-Onset Type 2 Diabetes (Y-T2D)
High metformin failure rates
Metformin response may be heritable
rs622342 (A>C) OCT1 variant
Fractional gluconeogenesis
Organic Cation Transporters (OCTs)
May affect metformin pharmacokinetics, fasting glucose, and gluconeogenesis
Randomized 3-month two-arm parallel trial
rs7903146 (G>T) TCF7L2 variant
Plasma glucose
[combination therapy only]
Metformin
Metformin + Liraglutide
Research Gap
Strongest glycemic predictors:
Baseline Glucose
Baseline Gluconeogenesis
Pharmacogenetics in Y-T2D are uncharacterized (OCT1 variants)
Serial pharmacokinetic sampling
Gluconeogenesis using ^2^H_2_O tracers
Pharmacogenetics → variable response in Youth-Onset Type 2 Diabetes
