## Supplemental Materials for "*OCT1* Variants Are Associated with Metformin Clearance and Gluconeogenesis: Mechanistic Insights for Youth-Onset Type 2 Diabetes in the MIGHTY Study"

### **Clinical Study Protocol**

#### **Study Agents/ Intervention**

Two study agents were used: metformin 500mg oral tablet and liraglutide (6mg/ml, 3ml) solution for subcutaneous injection, pre-filled, multi-dose pen that delivered doses of 0.6mg, 1.2mg, or 1.8mg. Both metformin and liraglutide were used within the approved dosing regimens. Neither drug was altered from the approved dosage formulation. Supplemental Table 3 illustrates the titration schedule for each study drug.

**Titration schedule for metformin and liraglutide**

| Study schedule | Week | Metformin  (oral tablet) | Liraglutide  (subcutaneous injection) |
| --- | --- | --- | --- |
| Run-in | -1 to 0 | - | - |
| After baseline visit | 0-1 | 500mg once daily | 0.6mg once daily |
|  | 1-2 | 500mg twice daily | 1.2mg once daily |
| **Target study dose** | 3-12 | 1000mg twice daily | 1.8mg once daily |

This table outlines the study drug titration schedule for participants receiving metformin (oral tablet) and liraglutide (subcutaneous injection). Doses were escalated weekly to reach the target study doses by Week 3.

Abbreviations: mg, milligram.

The participants started the study drug(s) on Day 2 of the baseline visit after the tracer protocol was completed. The Principal Investigator reviewed blood glucose logs and/ or CGM readings weekly and ensured that the study drug(s) were titrated to the highest tolerable dose, as guided by Table above. For patients on combination therapy, the dose of liraglutide was not increased if the fasting blood glucose was <75mg/dl on 2 or more consecutive days. If the fasting blood glucose subsequently rose to ≥90mg/dl on 2 or more consecutive days, the dose of liraglutide was increased by 0.6mg once daily every 5-7 days to achieve maximum tolerable dose.

#### **Dose titration for gastrointestinal intolerance**

For participants on the combination therapy arm who experienced gastrointestinal side effects, the dose of liraglutide was initially decreased by 0.6mg once daily. If gastrointestinal symptoms persisted after 2-3 days, the metformin dose was decreased by 500mg. If symptoms persisted, dose adjustments were made every 2-3 days, alternating between liraglutide (0.6mg) and metformin (500mg) dose reductions. If the highest tolerable metformin dose was < 1000mg daily or liraglutide <0.6mg daily, the subject was withdrawn from the study.

For participants on metformin alone, the dose of metformin was decreased by 500mg every 2-3 days until symptoms resolved. If the highest tolerable dose of metformin was <1000mg daily, the subject was withdrawn from the study.

If gastrointestinal symptoms resolved, and at the discretion of the PI, the dose of metformin and/or liraglutide was increased every 2-3 days to the maximum tolerable dose. Incremental dose increases were as follows: metformin 500mg every 2-3 days and liraglutide 0.6mg every 2-3 days (PMID: 37967247 and DOI: 10.6084/m9.figshare.24492187).

### **Supplemental Tables**

#### **Table 1:** Pharmacokinetic parameters by treatment groups

| **Parameter** | **Metformin (n=11)** | **Metformin + Liraglutide (n=8)** |
| --- | --- | --- |
| **Daily Metformin Dose: 1000 mg (500 mg BID)** | | |
|  | **N=1** | **N=2** |
| **CMAX (ng/mL)** | 956 | 607 (15%) |
| **TMAX (hr)^1^** | 2.1 | 3.1 (3.02-3.08) |
| **T1/2 (hr)** | 5.8 | 7.3 (68%) |
| **CMIN (ng/mL)** | 47.0 | 104 (55%) |
| **AUCTAU (hr*ng/mL)** | 7000 | 4596 (19%) |
| **CLSS/F (L/hr)** | 71.4 | 111 (18.6%) |
| **Daily Metformin Dose: 1500 mg (500 mg QAM, 1000 mg QPM)** | | |
|  | **N=2** | **N=2** |
| **CMAX (ng/mL)** | 1354 (29%) | 657 (91%) |
| **TMAX (hr)^1^** | 1.38 (1.13-1.62) | 5.1 (4.05-6.07) |
| **T1/2 (hr)** | 4.1 (14%) | 2.8 (12%) |
| **CMIN (ng/mL)** | 498 (32%) | 88.3 (99%) |
| **AUCTAU (hr*ng/mL)** | 8764 (28%) | 4281 (87%) |
| **CLSS/F (L/hr)** | 59.3 (27.6%) | 187 (86.7%) |
| **Daily Metformin Dose: 2000 mg (1000 mg BID)** | | |
|  | **N=8** | **N=4** |
| **CMAX (ng/mL)** | 1777 (32.5%) | 1297 (43%) |
| **TMAX (hr)^1^** | 3.0 (1.53-4.02) | 2.3 (1.37-8.03) |
| **T1/2 (hr)** | 5.1 (88.7%) | 3.5 (29%)^2^ |
| **CMIN (ng/mL)** | 446 (55%) | 227 (60%) |
| **AUCTAU (hr*ng/mL)** | 11101 (34.2%) | 8013 (17%) |
| **CLSS/F (L/hr)** | 98.9 (31%) | 126 (23%)^2^ |

This table summarizes steady-state pharmacokinetic parameters for metformin alone and in combination with liraglutide, stratified by daily metformin dose. The values presented are arithmetic means (% coefficient of variation, %CV) unless otherwise specified.

^1^ T_MAX_ reported as median [range]; 2 n=3 as one patient did not meet Λ_Z_ acceptance criteria

Abbreviations: CMAX, maximum plasma concentration; TMAX, time to maximum plasma concentration; T1/2, elimination half-life; CMIN, trough concentration; AUCTAU, area under the concentration–time curve during the dosing interval; CLSS/F, clearance at steady state; %CV, percent coefficient of variation; BID, twice daily; QAM, every morning; QPM, every evening.

#### **Table 2:** Baseline Participant Characteristics by Group

|  | **Diet & Lifestyle**  **(n=5)** | **MET**  **(n=14)** | **MET+LIRA**  **(n=11)** |
| --- | --- | --- | --- |
| **Age (years)** | 16.6 ± 3.6 | 15.6 ± 2.1 | 15 ± 2.1 |
| **Female sex n (%)** | 2 (40) | 7 (50) | 7 (64) |
| **Time since diagnosis (years)** | 1.73 | 2.14 | 1.53 |
| **Metformin naïve n (%)** | 5 (100) | 1 (7) | 5 (45) |
| **Systolic Blood Pressure (mmHg)** | 131 ± 10 | 131 ± 14 | 128 ± 14 |
| **Weight (kg)** | 108.2 ± 32.3 | 116.2 ± 25.3 | 113.3 ± 23.3 |
| **Body Mass Index (kg/m^2^)** | 38.3 ± 8.3 | 39.4 ± 7.9 | 38.2 ± 8.0 |
| **Lean body mass (kg)** | 50.7 ± 13.2 | 55.7 ± 9.8 | 56.4 ± 9.6 |
| **Hemoglobin A1c (%)** | 5.7 ± 0.3 | 6.4 ± 0.7 | 7.4 ± 0.9 |
| **Fasting glucose (mg/dL)** | 111 ± 19 | 119 ± 26 | 142 ± 48 |
| **2-hour glucose (mg/dL)** | 192 ± 82 | 239 ± 72  (n=13) | 273 ± 88 |
| **eGFR (Bedside Schwartz, mL/min/1.73 m²)** | 93 ± 4 | 112 ± 27 | 113 ± 19 |
| **eGFR (CKiD U25, mL/min/1.73 m²)** | 95 ± 15 | 115 ± 31 | 116 ± 25 |
| **Serum creatinine (mg/dL)** | 0.7 ± 0.1 | 0.7 ± 0.2 | 0.7 ± 0.2 |
| **Fractional Gluconeogenesis (%)** | — | 67 ± 13  (n=11) | 66 ± 10  (n=11) |
| **Metformin Genetic Risk Score** | 3.1 ± 0.8 | 3.8 ± 1.1 | 3.1 ± 0.8 |

This table summarizes the demographic and clinical characteristics of study participants at baseline. Data are presented as mean ± standard deviation (SD), *n* (%), or median (interquartile range, IQR), as appropriate. Abbreviations: MET, metformin; MET+LIRA, Metformin-Liraglutide; eGFR, estimated glomerular filtration rate

#### **Table 3.** Baseline characteristics for participants who completed pharmacokinetic sampling

|  | **Pre-Met (n=11)** | **Pre-Met+Lira (n=8)** |
| --- | --- | --- |
| **Age (years)** | 15.36 ± 2.16 | 15.13 ± 2.47 |
| **Female sex n (%)** | 7 (64) | 6 (75) |
| **Time since diagnosis (years)** | 2.12 ± 1.52 | 1.56 ± 1.04 |
| **Metformin naive n (%)** | 10 (91) | 4 (50) |
| **Systolic Blood Pressure (mmHg)** | 129.7 ± 14.5 | 128.9 ± 15.8 |
| **Weight (kg)** | 115.8 ± 25.2 | 115.2 ± 27.5 |
| **BMI (kg/m^2^)** | 40.8 ± 7.7 | 39.1 ± 9.3 |
| **Lean body mass (kg)** | 54.4 ± 9.4 | 56.0 ± 11.3 |
| **HbA1c (%)** | 6.4 ± 0.7 | 7.2 ± 0.8 |
| **Fasting glucose (mg/dL)** | 116.6 ± 26.9 | 125.5 ± 29.9 |
| **2-hour glucose (mg/dL)** | 227.3 ± 65.5 | 243.5 ± 61.6 |
| **Fractional Gluconeogenesis (%)** | 67.2 ± 13.1 | 64.5 ± 11.1 |
| **Serum creatinine (mg/dL)** | 0.62 ± 0.13 | 0.67 ± 0.18 |
| **eGFR (Bedside Schwartz, mL/min/1.73 m²)** | 116.7 ± 24.9 | 110.7 ± 18.7 |
| **eGFR (CKiD U25, mL/min/1.73 m²)** | 121.6 ± 29.0 | 115.7 ± 26.0 |
| **Genetic Risk Score** | 3.7 ± 1.2 | 3.4 ± 0.8 |

This table summarizes the demographic and clinical characteristics of study participants at baseline who completed pharmacokinetic sampling. Participants are stratified by treatment group allocation and variables assessed are age, sex, diagnosis, time since diagnosis, metformin naïve, systolic blood pressure, weight, body mass index, lean body mass, hemoglobin A1c, fasting glucose, 2-hour glucose, eGFR (estimated glomerular filtration rate), serum creatinine, and fractional gluconeogenesis.

Data are presented as mean ± standard deviation (SD), *n* (%), or median (interquartile range, IQR), as appropriate.

Analysis of variance (ANOVA) was used for comparisons of continuous variables across groups, and Fisher’s exact test was used for categorical variables

Abbreviations: eGFR, estimated glomerular filtration rate

#### **Table 4:** Effect of type 2 diabetes risk variant rs7903146 *TCF7L2* on glycemia

| **Model Metrics** | **Post-Treatment Steady State Fasting Plasma Glucose (mmol/L)** | |
| --- | --- | --- |
|  | **β (95% CI)** | **Variable *P*-value** |
| **rs7903146_*TCF7L2* [All Subjects, n=21]** | 0.25 (-0.56, 1.07) | 0.512 |
| **Drug** | 0.32 (-0.86, 1.50) | 0.572 |
| **Drug x rs7903146_*TCF7L2*** | -1.33 (-2.73, 0.07) | 0.060 |
| **Baseline Fasting Plasma Glucose** | 3.73 (-0.30, 7.76) | 0.067 |
| **Age** | -0.02 (-0.14, 0.10) | 0.779 |
| **Sex** | 0.50 (-0.16, 1.15) | 0.127 |
|  | **Adjusted R^2^:** 0.609, **Model *P*-Value:** 0.002 | |
| **rs7903146_*TCF7L2* [MET+LIRA, n=10]** | -1.32 (-2.42, -0.22) | 0.028 |
| **Baseline FPG** | 2.98 (-2.39, 8.34) | 0.213 |
| **Age** | -0.04 (-0.24, 0.16) | 0.642 |
| **Sex** | 0.16 (-0.83, 1.15) | 0.698 |
|  | **Adjusted R^2^:** 0.776, **Model *P*-Value:** 0.018 | |

This table presents linear regression models evaluating the association of the *TCF7L2* variant *rs7903146* with steady-state fasting plasma glucose in youth treated with metformin or metformin + liraglutide. Ordinary least squares regression was performed for the overall cohort (n = 21), including a genotype-by-treatment interaction term to account for unequal distribution of rs7903146 across treatment groups. A separate model was fit for the metformin plus liraglutide–treated subgroup (MET+LIRA; n = 10). Each model reports the β coefficient (95% confidence interval) and corresponding *P*-value for each covariate. Model fit is indicated by the adjusted R² and overall model *P*-value.

Abbreviations: *TCF7L2*, transcription factor 7-like 2; FPG, fasting plasma glucose; β, regression coefficient; CI, confidence interval, MET+LIRA, metformin + liraglutide.

#### **Table 5:** Summary of variants evaluated for pharmacogenetic effects

| **dbSNP ID** | **Nearest Gene** | **Chr:bp^a^** | **Risk/Other Allele** | **Risk Allele Frequency** | **Risk Allele Frequency (ASW)*** | **Risk Allele Frequency (EUR)**** | **Variant Coding** | **Reported Association(s)** | **Ref^#^** |
| --- | --- | --- | --- | --- | --- | --- | --- | --- | --- |
| **Metformin Variants** | | | | | | | | | |
| rs628031 | *OCT1* | 6:160139813 | G/A | 0.82 | 0.71 | 0.59 | **Additive** | ↑ Met intolerance  ↑ HbA1c | (1-3) |
| rs622342 | *OCT1* | 6:160151834 | C/A | 0.22 | 0.21 | 0.38 | **Additive** | ↑ HbA1c | (3-9) |
| rs461473 | *OCT1* | 6:160122530 | G/A | 0.95 | 0.95 | 0.91 | **Dominant** | ↑ HbA1c | (10) |
| rs662301 | *OCT2* | 6:160275887 | T/C | 0.02 | 0.02 | 0.07 | **Additive** | ↓ T2D incidence | (11, 12) |
| rs316019 | *OCT2* | 6:160249250 | C/A | 0.9 | 0.85 | 0.89 | **Recessive** | ↓ HbA1c | (13, 14) |
| rs8192675 | *GLUT2* | 3:171007094 | T/C | 0.25 | 0.35 | 0.62 | **Additive** | ↓ HbA1c | (15-18) |
| rs8065082 | *MATE1* | 17:19561878 | C/T | 0.67 | 0.71 | 0.54 | **Additive** | ↓ T2D incidence | (11) |
| rs2252281 | *MATE1* | 17:19533874 | C/T | ‡ | ‡ | 0.29 | **Dominant** | ↓ HbA1c | (19) |
| rs12943590 | *MATE2* | 17:19716685 | A/G | 0.25 | 0.22 | 0.27 | **Dominant** | ↑ Met clearance  ↑ HbA1c | (8, 14, 19-22) |
| rs11212617 | *ATM* | 11:108412434 | A/C | 0.33 | 0.37 | 0.41 | **Additive** | ↑ HbA1c | (8, 10, 23-26) |
| **Liraglutide Variants** | | | | | | | | | |
| rs7903146 | *TCF7L2* | 10:112998590 | T/C | 0.45 | 0.36 | 0.32 | **Dominant** | ↓ Weight (Lira)  ± Weight (Met) | (27-29) |
| rs6923761 | *GLP-1R* | 6:39066296 | G/A | 0.98 | 0.92 | 0.67 | **Recessive** | ↓ % Total fat | (27, 28, 30, 31) |
| rs7202877 | *CTRB1/2* | 16:75213347 | T/G | 0.83 | 0.85 | 0.90 | **Dominant** | ↑ Insulin secretion | (32, 33) |

This table summarizes key genetic variants associated with modulation of glycemia or MET/LIRA treatment response and includes additional references for reported associations. Variants were selected based on effects observed in literature and stratified by predicted effects on metformin or liraglutide treatment. Study participant genetic frequencies were compared against the 1000 Genomes Project (34). Each entry lists the dbSNP identifier, nearest gene, chromosomal location (Chr:bp) ^a^, risk allele, risk allele frequency, risk allele frequency in the 1000 Genomes populations for ASW (African Ancestry individuals in the U.S. Southwest) ^b^, risk allele frequency in the 1000 Genomes EUR population (European Ancestry individuals) ^c^, variant coding used in analyses, and reported pharmacogenetic associations.

^a^ Chromosome and base-pair position based on GRCh38.

^b^ Risk allele frequency in the 1000 Genomes ASW population (African Ancestry individuals in the U.S. Southwest)

^c^ Risk allele frequency in the 1000 Genomes EUR population (European Ancestry individuals)

‡ The genotype for this variant was not available in this study or the 1000 Genomes Project.

Abbreviations: ASW, African Ancestry individuals in the U.S. Southwest HbA1c; AFR, African Ancestry individuals, EUR, European Ancestry individual; HbA1c, hemoglobin A1c; Met, metformin; Lira, liraglutide; Ref, reference

### **Supplemental Figures**

#### **Supplemental Figure 1: Participant flow diagram**

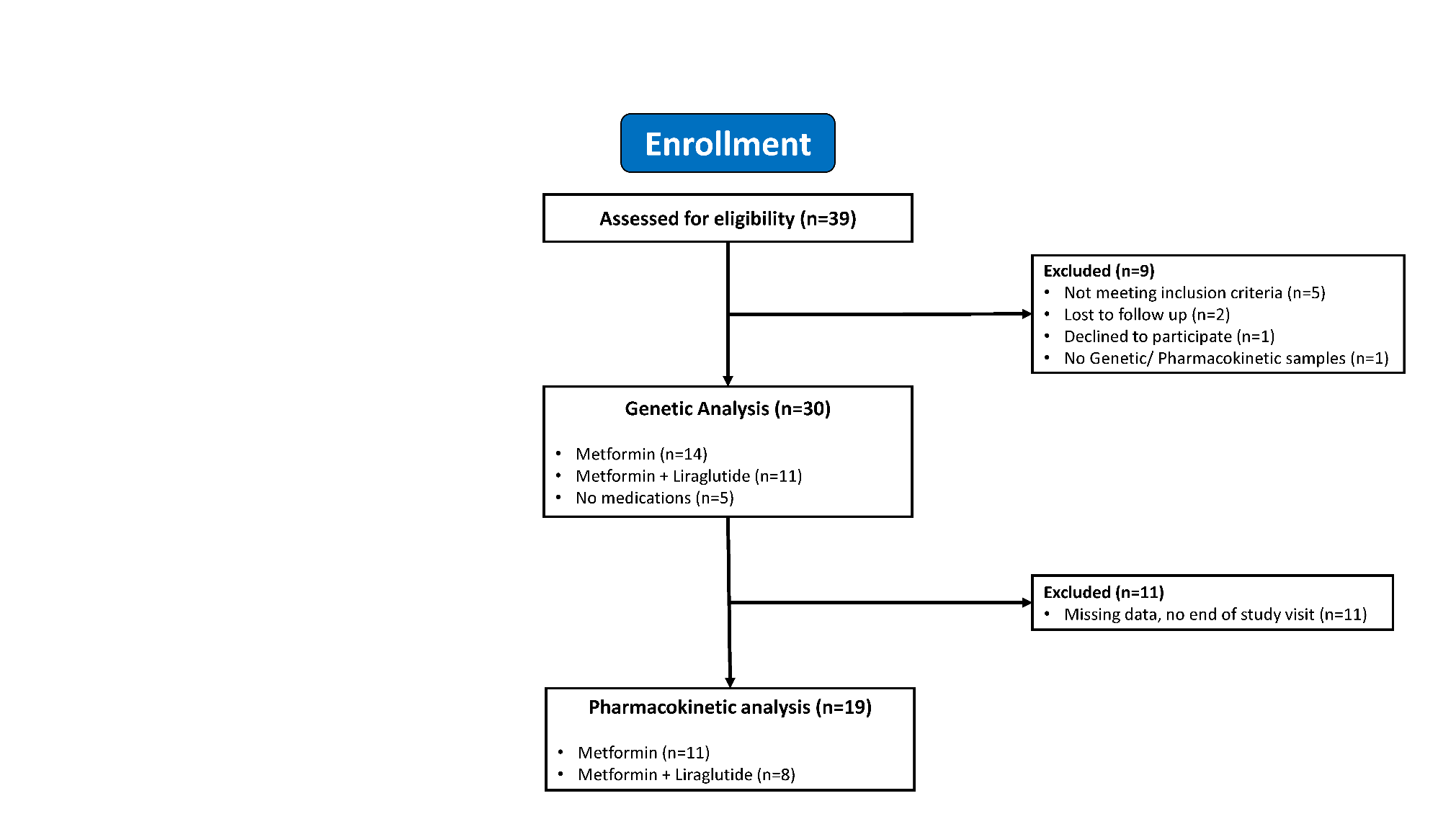

This figure shows the flow of participants through screening, enrollment, and analysis. Of the 39 individuals assessed for eligibility, 30 were included in the genetic analysis cohort and 19 in the pharmacokinetic analysis cohort. The primary inclusion criterion was study completion, genetic testing, and pharmacokinetic sampling. Participants were excluded for not meeting inclusion criteria, loss to follow-up, withdrawal of consent, or missing biological sample

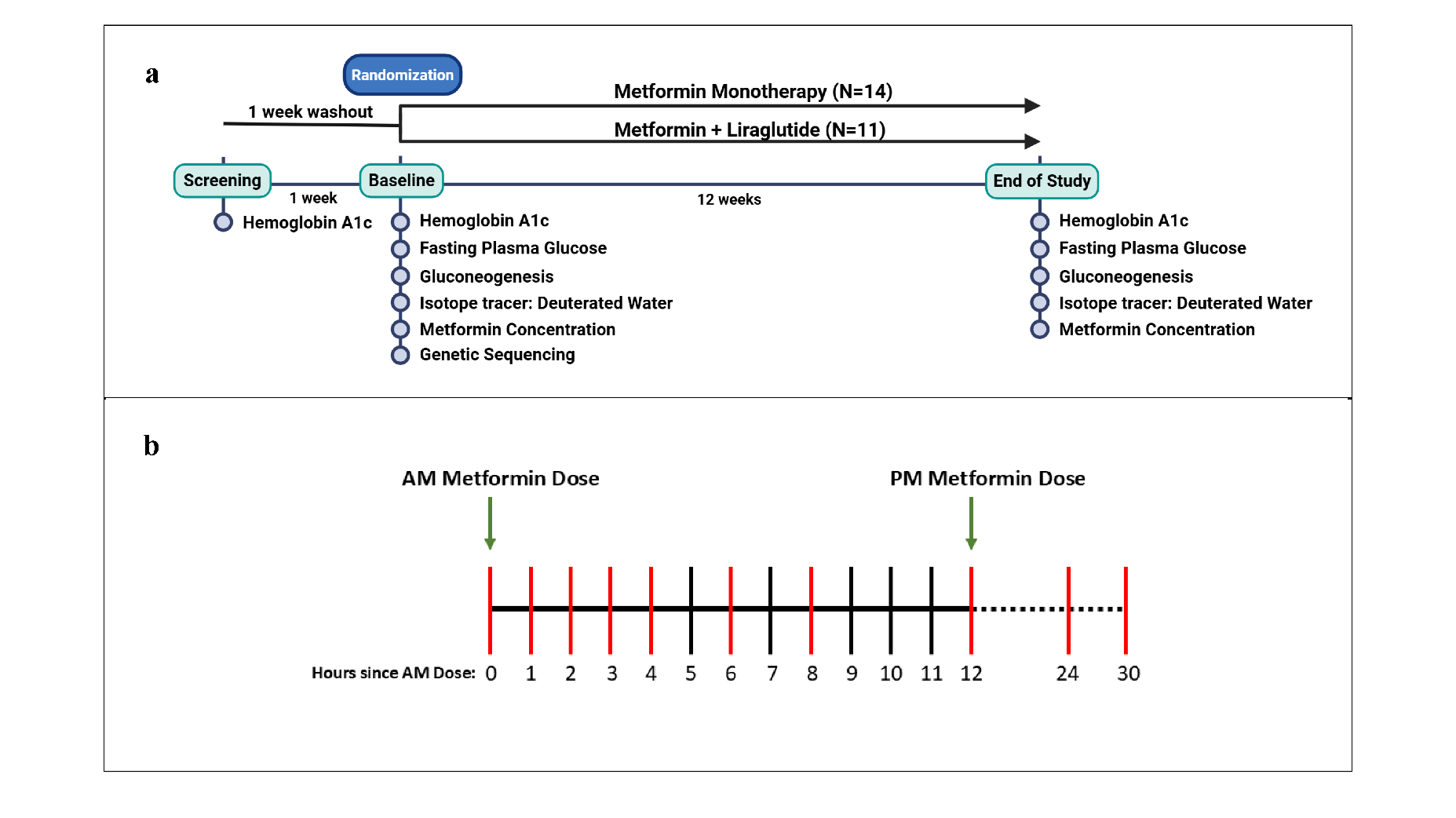

#### **Supplemental Figure 2: (a-b): Study design and metformin pharmacokinetic sampling schedule**

These panels describe the study design and serial metformin pharmacokinetic blood plasma sampling. Panel **(a)** displays the overall study design showing screening, baseline, and end-of-study visits. Randomization of intervention occurred at the baseline visit and participants were assigned either metformin monotherapy or metformin and liraglutide therapy. At baseline visit and end of study participants underwent testing of Hemoglobin A1c (HbA1c), fasting plasma glucose, gluconeogenesis (measured with deuterated water isotope tracer), and serial metformin pharmacokinetic blood sampling. Genetic sequencing occurred only at baseline. Panel **(b)** illustrates the study schedule for timing in hours of pharmacokinetic blood draws during baseline and end-of-study visits. Green arrows indicate timing of metformin administration in morning (AM) and evening (PM) doses. The red bars indicate timing of blood plasma PK sampling during visits. Created with BioRender.com.

Abbreviations: AM, morning; PM, evening.

#### **Supplemental Figure 3: Observed and model-predicted red blood cell metformin concentrations**

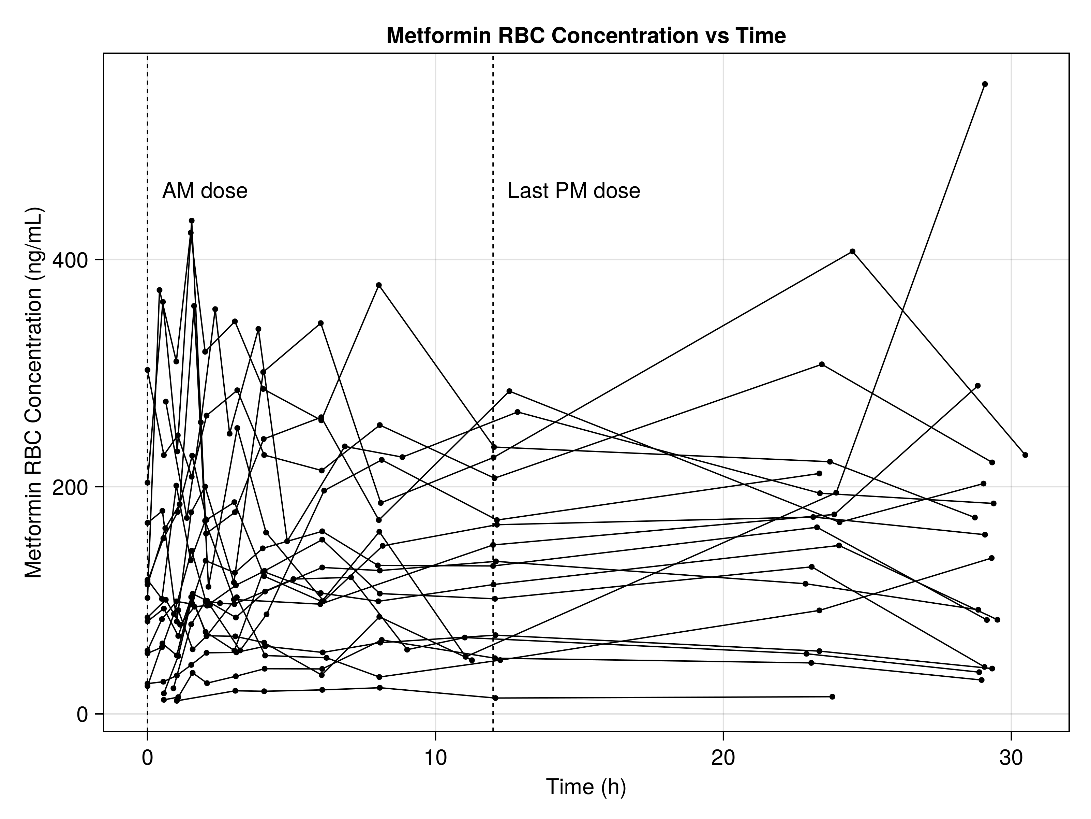

This figure shows the observed metformin concentrations in red blood cells over time for each participant that underwent pharmacokinetic testing. As shown in the figure, these values were highly variable.

Abbreviations: RBC, Red Blood Cell
